# Discovery and systematic characterization of risk variants and genes for coronary artery disease in over a million participants

**DOI:** 10.1101/2021.05.24.21257377

**Authors:** Krishna G Aragam, Tao Jiang, Anuj Goel, Stavroula Kanoni, Brooke N Wolford, Elle M Weeks, Minxian Wang, George Hindy, Wei Zhou, Christopher Grace, Carolina Roselli, Nicholas A Marston, Frederick K Kamanu, Ida Surakka, Loreto Muñoz Venegas, Paul Sherliker, Satoshi Koyama, Kazuyoshi Ishigaki, Bjørn O Åsvold, Michael R Brown, Ben Brumpton, Paul S de Vries, Olga Giannakopoulou, Panagiota Giardoglou, Daniel F Gudbjartsson, Ulrich Güldener, Syed M. Ijlal Haider, Anna Helgadottir, Maysson Ibrahim, Adnan Kastrati, Thorsten Kessler, Ling Li, Lijiang Ma, Thomas Meitinger, Sören Mucha, Matthias Munz, Federico Murgia, Jonas B Nielsen, Markus M Nöthen, Shichao Pang, Tobias Reinberger, Gudmar Thorleifsson, Moritz von Scheidt, Jacob K Ulirsch, EPIC-CVD Consortium, Biobank Japan, David O Arnar, Deepak S Atri, Noël P Burtt, Maria C Costanzo, Jason Flannick, Rajat M Gupta, Kaoru Ito, Dong-Keun Jang, Yoichiro Kamatani, Amit V Khera, Issei Komuro, Iftikhar J Kullo, Luca A Lotta, Christopher P Nelson, Robert Roberts, Gudmundur Thorgeirsson, Unnur Thorsteinsdottir, Thomas R Webb, Aris Baras, Johan LM Björkegren, Eric Boerwinkle, George Dedoussis, Hilma Holm, Kristian Hveem, Olle Melander, Alanna C Morrison, Marju Orho-Melander, Loukianos S Rallidis, Arno Ruusalepp, Marc S Sabatine, Kari Stefansson, Pierre Zalloua, Patrick T Ellinor, Martin Farrall, John Danesh, Christian T Ruff, Hilary K Finucane, Jemma C Hopewell, Robert Clarke, Jeanette Erdmann, Nilesh J Samani, Heribert Schunkert, Hugh Watkins, Cristen J Willer, Panos Deloukas, Sekar Kathiresan, Adam S Butterworth, on behalf of the CARDIoGRAMplusC4D Consortium

## Abstract

Rapid progress of the discovery of genetic loci associated with common, complex diseases has outpaced the elucidation of mechanisms pertinent to disease pathogenesis. To address relevant barriers for coronary artery disease (CAD), we combined genetic discovery analyses with downstream characterization of likely causal variants, genes, and biological pathways. Specifically, we conducted a genome-wide association study (GWAS) comprising 181,522 cases of CAD among 1,165,690 participants. We detected 241 associations, including 54 associations and 30 loci not previously linked to CAD. Next, we prioritized likely causal variants using functionally-informed fine-mapping, yielding 42 associations with fewer than five variants in the 95% credible set. Combining eight complementary predictors, we prioritized 185 candidate causal genes, including 94 genes supported by three or more predictors. Similarity-based clustering underscored a role for early developmental processes, cell cycle signaling, and vascular proliferation in the pathogenesis of CAD. Our analysis identifies and systematically characterizes risk loci for CAD to inform experimental interrogation of putative causal mechanisms for CAD.

## INTRODUCTION

Coronary artery disease (CAD) remains the leading global cause of mortality, principally reflecting effects of risk behaviors and genetic susceptibility.[1] Previous genetic association studies have identified over 200 susceptibility loci for CAD. Consistent with other common, complex diseases, genetic discovery analyses have identified the polygenic architecture of CAD, enabled insights into disease etiology and causal risk factors, and facilitated the development of novel tools for clinical risk prediction.[2-10] However, with rapid increases in the availability of large-scale human genetic data linked to health outcomes, the identification of disease-associated genetic loci has outpaced their ensuing functional characterization.

Several *in silico* tools have emerged to help determine the mechanisms connecting regions of the genome to disease risk.[11, 12] Nonetheless, it remains fundamentally challenging to identify the causal genes underlying genetic associations as these tools can produce spurious findings and frequently lack consensus.[13] Recent analyses have suggested the value of integrating locus-specific (“locus-based”) approaches to gene prioritization with more global (“similarity-based”) assessments of shared pathways and functions to enhance the prediction of putative causal genes.[13-15] The integration of multiple orthogonal lines of evidence, and the use of disease-specific resources to aid variant and gene classifications, may expedite the transition from gene maps to disease mechanisms.

To extend these approaches to CAD, we first analyzed imputed genotyping array data from ten studies, comprising over 120,000 cases of CAD and 700,000 controls. We then combined these results with summary statistics from the CARDIoGRAMplusC4D Consortium, achieving a total sample of 181,522 CAD cases among 1,165,690 study participants.[2, 7, 10, 16] Our primary objectives were to: (1) discover novel genetic associations with CAD; (2) determine the impact of expanded genetic discovery for identifying loci of biological relevance and improving clinical risk prediction; and (3) implement a systematic and integrative approach – including well-established and newer methods – to prioritize likely causal variants and genes at genome-wide significant associations for CAD, thereby providing a catalogue of high-priority testable hypotheses for experimental follow-up.

## RESULTS

### Discovery of known and novel CAD loci

Participants were largely (>95%) of European ancestry (predominantly from Europe or the US) and 46% were female (<u>Supplementary Table 1</u>). After quality control and filtering, 20,073,070 variants were included in the discovery meta-analysis (<u>Online Methods</u>). To identify independent variants, we performed approximate conditional analysis using GCTA-COJO, and report 241 independently associated variants that exceeded genome-wide significance (p-value≤5.0×10^−8^) at 198 loci (<u>Supplementary Table 2</u>; <u>Supplementary Figure 1</u>). 54 sentinel variants were uncorrelated (r^2^ <0.2) with variants reported in previous large-scale genetic analyses, including 30 that lie outside genomic regions previously reported for CAD (**Table 1**). A phenome-wide association scan (PheWAS) in UK Biobank indicated that 130 (54%) of the 241 CAD-associated variants were not associated (p-value>3.9×10^−6^) with conventional CAD risk factors such as blood lipids, blood pressure, type 2 diabetes, or adiposity (<u>Supplementary Table 3</u>), suggesting widespread mediation of CAD risk via other mechanisms.

**Table 1.** 30 novel loci for CAD.

| Nearest gene | Lead variant rsID | Chr | Position | Effect allele | Non-effect allele | Odds ratio | 95% CI | P-value |
| --- | --- | --- | --- | --- | --- | --- | --- | --- |
| *KDF1* | rs79598313 | 1 | 27,284,913 | T | C | 1.10 | (1.06-1.14) | 3.6x10<sup>-8</sup> |
| *LOC100131060* | rs71646019 | 1 | 59,433,354 | T | C | 1.04 | (1.03-1.05) | 6.1x10<sup>-10</sup> |
| *OTUD7B* | rs67807996 | 1 | 149,995,265 | A | G | 1.04 | (1.03-1.05) | 1.1x10<sup>-12</sup> |
| *MIR4432* | rs243071 | 2 | 60,619,028 | A | G | 1.03 | (1.02-1.04) | 2.7x10<sup>-8</sup> |
| *SAP130* | rs114192718 | 2 | 128,785,663 | T | C | 1.06 | (1.04-1.08) | 2.6x10<sup>-8</sup> |
| *ACVR2A* | rs35611688 | 2 | 148,377,860 | T | C | 0.97 | (0.96-0.98) | 1.5x10<sup>-8</sup> |
| *LNX1* | rs17083333 | 4 | 54,572,066 | T | G | 0.97 | (0.96-0.98) | 1.2x10<sup>-8</sup> |
| *ITGA1* | rs4074793 | 5 | 52,193,125 | A | G | 0.95 | (0.93-0.97) | 1.6x10<sup>-8</sup> |
| *FER* | rs112949822 | 5 | 108,085,190 | A | G | 0.95 | (0.93-0.96) | 1.1x10<sup>-9</sup> |
| *DMXL1* | rs13169691 | 5 | 118,448,279 | T | C | 1.04 | (1.03-1.06) | 2.6x10<sup>-8</sup> |
| *FBN2* | rs6883598 | 5 | 127,926,190 | A | C | 0.97 | (0.96-0.98) | 9.7x10<sup>-10</sup> |
| *PTK7* | rs1034246 | 6 | 43,068,370 | T | G | 0.97 | (0.96-0.98) | 6.4x10<sup>-10</sup> |
| *MACC1* | rs10486389 | 7 | 20,300,416 | A | G | 0.97 | (0.96-0.98) | 6.5x10<sup>-10</sup> |
| *C9orf146* | rs10961206 | 9 | 13,724,051 | A | T | 1.05 | (1.04-1.07) | 8.1x10<sup>-10</sup> |
| *ACER2* | rs10811183 | 9 | 19,436,055 | A | G | 1.04 | (1.02-1.05) | 1.6x10<sup>-8</sup> |
| *C5* | rs41312891 | 9 | 123,726,749 | G | GCAAA | 0.94 | (0.92-0.96) | 5.9x10<sup>-9</sup> |
| *PLCE1* | rs55753709 | 10 | 96,029,170 | T | C | 0.96 | (0.95-0.97) | 2.2x10<sup>-13</sup> |
| *R3HCC1L* | rs884811 | 10 | 99,923,763 | C | G | 1.03 | (1.02-1.04) | 3.1x10<sup>-9</sup> |
| *MMP13* | rs1892971 | 11 | 102,795,606 | A | G | 0.96 | (0.95-0.97) | 5.1x10<sup>-10</sup> |
| *ST3GAL4* | rs10790800 | 11 | 126,262,638 | A | G | 1.03 | (1.02-1.04) | 9.1x10<sup>-9</sup> |
| *TBX3* | rs34606058 | 12 | 115,353,368 | T | C | 0.97 | (0.96-0.98) | 7.7x10<sup>-9</sup> |
| *DOCK9* | rs8000794 | 13 | 99,434,810 | C | G | 1.03 | (1.02-1.04) | 4.3x10<sup>-8</sup> |
| *LIPC* | rs588136 | 15 | 58,730,498 | T | C | 0.96 | (0.95-0.98) | 7.0x10<sup>-10</sup> |
| *UNC13D* | rs2410859 | 17 | 73,841,285 | T | C | 1.03 | (1.02-1.04) | 4.3x10<sup>-9</sup> |
| *CPLX4* | rs11663411 | 18 | 56,960,510 | T | C | 0.97 | (0.96-0.98) | 2.6x10<sup>-8</sup> |
| *MYO9B* | rs7246865 | 19 | 17,219,105 | A | G | 1.03 | (1.02-1.05) | 1.9x10<sup>-8</sup> |
| *RRBP1* | rs1132274 | 20 | 17,596,155 | A | C | 1.04 | (1.03-1.05) | 1.8x10<sup>-8</sup> |
| *MAFB* | rs2207132 | 20 | 39,142,516 | A | G | 1.10 | (1.07-1.13) | 6.7x10<sup>-10</sup> |
| *ARVCF* | rs71313931 | 22 | 19,960,184 | C | G | 0.97 | (0.96-0.98) | 2.3x10<sup>-9</sup> |
| *SCUBE1* | rs139012 | 22 | 43,623,972 | A | G | 0.97 | (0.96-0.98) | 2.1x10<sup>-8</sup> |

Positions are according to GRCh37. Odds ratios (and 95% confidence intervals) are for per-allele effect estimates according to the effect allele.

Several of the novel associations (**Table 1**) were found near mechanistically plausible causal genes, including: rs35611688 near *ACVR2A*, which encodes a receptor for activin A, a member of the transforming growth factor (TGF)-beta superfamily of cytokines implicated in atherogenesis;[17-19] rs6883598 near *FBN2*, encoding fibrillin-2, which mediates the early stages of elastic fiber assembly and is associated with aortic aneurysms and Beals Syndrome, a Marfan-like disorder;[20-22] and rs1892971 near *MMP13*, which encodes matrix metalloproteinase (MMP)-13, an interstitial collagenase that influences the structural integrity of atherosclerotic plaques through regulation and organization of intraplaque collagen.[23, 24] While the sentinel variant near *FBN2* was associated with blood pressure and hypertension in the PheWAS, the lead variants near *ACVR2A* and *MMP13* were not associated with conventional CAD risk factors, suggesting they are likely to act through alternative pathways.

### Allelic architecture

Of the 54 novel associations, 46 sentinel variants were common (minor allele frequency [MAF]>0.05) with relatively weak effects on CAD (odds ratio [OR] per CAD risk allele from 1.03-1.07) (**Figure 1**). The remaining eight were low-frequency (MAF=0.009 to 0.036), of which four had comparatively strong effects (OR=1.30 to 1.44) and four had more modest effect associations (OR=1.10 to 1.14) (<u>Supplementary Figure 2</u>). To boost power to detect associations driven by rarer variants, we conducted gene-based tests of missense and predicted loss-of-function variants in UK Biobank (n=33,941 CAD cases, 438,394 controls; <u>Supplementary Table 4</u>). Apart from a strong signal for *PCSK9*, we did not find evidence for further association with a burden of low-frequency or rare variants (<u>Supplementary Figure 3</u>; <u>Supplementary Table 5</u>).

**Figure 1.**
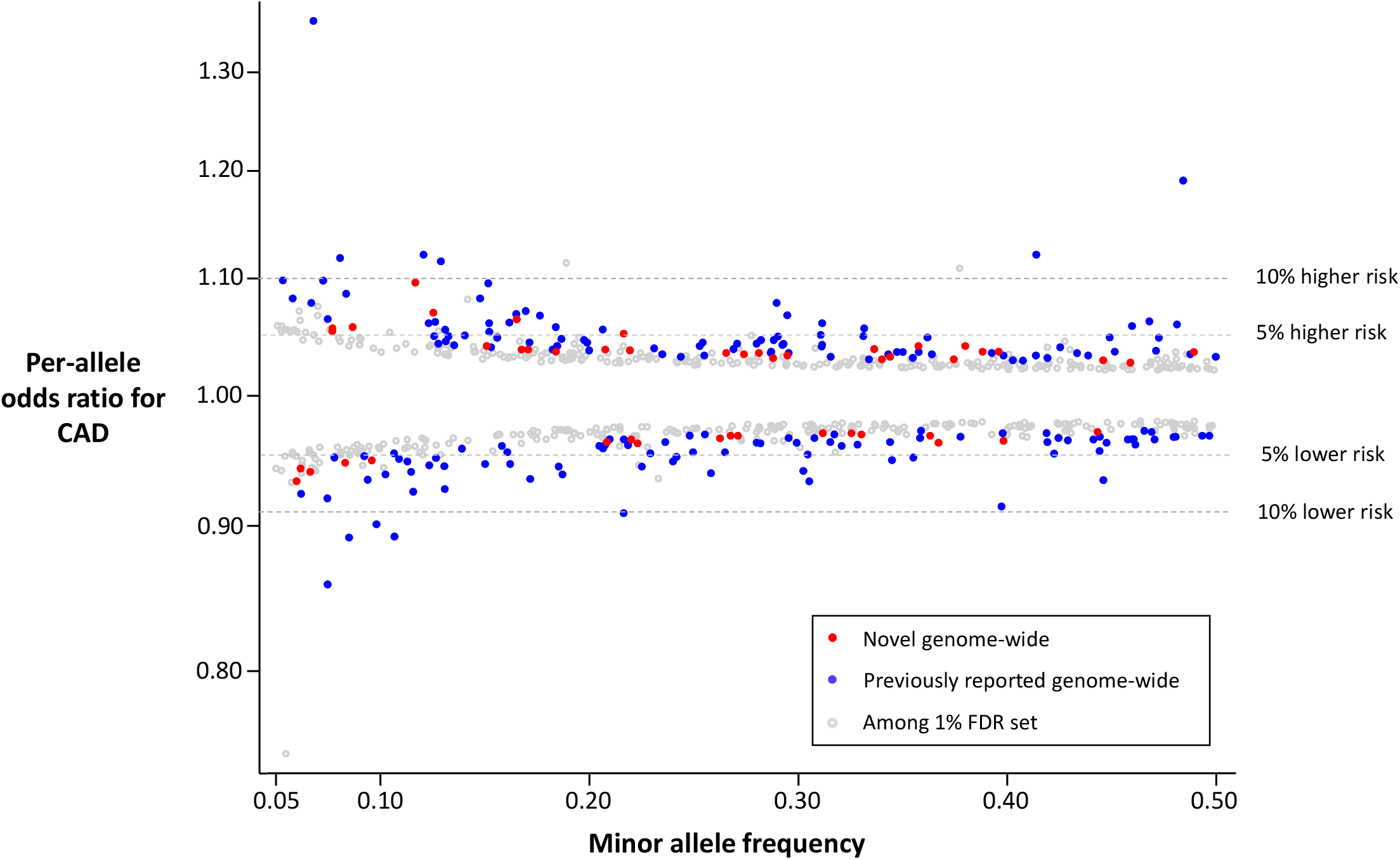
Common variant association signals for CAD. **Minor allele frequency versus per-allele odds ratio for CAD for common sentinel variant (MAF>5%) associations reaching genome-wide significance or the 1% FDR threshold in our study**. Colored circles indicate genome-wide significant associations (p-value<5.0×10^−8^) with sentinel variants that are not correlated (r^2^ <0.2) with a previously reported variant (‘novel’ – red), genome-wide significant sentinel variants correlated with a previously reported variant (‘known’ - blue), and associations reaching the 1% FDR threshold (p-value<2.52×10^−5^) in our meta-analysis (grey).

### Differential effects by sex

To identify associations that differ by sex, we conducted sex-stratified GWAS in a subset of 16 studies comprising 77,080 CAD cases (<u>Supplementary Table 6</u>). After combining results across studies using a sex-differentiated meta-analysis, which allows for between-sex heterogeneity, we found ten associations (nine previously reported) that reached genome-wide significance (p-value≤5.0×10^−8^) and had evidence (p-value≤0.01) for between-sex heterogeneity (<u>Supplementary Table 7</u>). Nine of these had stronger effects in the male-only analysis - including associations at the well-known 9p21 and *SORT1* loci - however rs7696877 near *MYOZ2* had a stronger effect in females (per-allele OR=0.94) than males (per-allele OR=0.98; heterogeneity p-value=0.007).

### Sub-threshold associations

At a significance level (p-value<2.52×10^−5^) approximating a 1% false discovery rate (FDR), we identified a further 47,622 variants associated with CAD, including 656 conditionally independent associations (<u>Supplementary Table 8</u>). The majority (486, 74.1%) were common variants (MAF>0.05), but almost all had relatively weak effects (per-allele OR<1.07). Among these were several associations with strong biological priors, including rs41279633 (p-value=1.24×10^−6^) in *NPC1L1*, which encodes Niemann-Pick C1-like 1, an important mediator of intestinal cholesterol absorption and the target of ezetimibe, a cholesterol lowering drug. Other examples include loci known to be associated with cardiovascular risk factors, such as *PNPLA3* (rs738408; p-value=1.04×10^−5^), the strongest locus for non-alcoholic fatty liver disease[25], and *TCF7L2* (rs7903146; p-value=6.39×10^−8^), the strongest locus for type 2 diabetes[26]. Heritability for liability to CAD was estimated to be 15.5% for the 241 conditionally independent associations reaching genome-wide significance, increasing to 36.1% for the 897 associations with p-value<2.52×10^−5^.

### Trans-ethnic comparison and meta-analysis

The recent publication of a large GWAS from Biobank Japan permitted evaluation of the genome-wide associations in a well-powered set of East Asian ancestry participants.[3] Effect estimates for the 199 sentinel variants in both datasets were strongly positively correlated (*r*=0.59) between the predominantly European ancestry meta-analysis and the Biobank Japan GWAS (<u>Supplementary Figure 4a</u>), as were the effect allele frequencies (*r*=0.76; <u>Supplementary Figure 4b</u>). To assess the potential for enhanced discovery by combining results from different ethnic groups, we then meta-analyzed the Biobank Japan GWAS summary statistics with those from the current analysis, yielding 38 additional novel loci at genome-wide significance (<u>Supplementary Table 9</u>). 36 of these were included in the 1% FDR set, including the aforementioned associations at *TCF7L2* and *PNPLA3*. The exceptions were two variants (rs5867305 in *SKP2* and rs75655731 near *LINC00599*) that are considerably more common in East Asians and had stronger effect estimates in Biobank Japan (<u>Supplementary Table 9</u>).

### Association of polygenic risk scores with incident and recurrent CAD

To assess the impact of the enhanced discovery sample size on genetic risk prediction for CAD, we constructed and evaluated 362 polygenic risk scores (PRS) using combinations of PRS derivation methods (Pruning and Thresholding[27] or LDpred algorithm[28]) and summary statistics from either the current meta-analysis or a 1000 Genomes-imputed GWAS involving around 60,000 CAD cases published in 2015.[7] We selected the optimized PRS for each combination of derivation method and GWAS summary statistics based on performance when predicting incident CAD in a training dataset from the Malmo Diet and Cancer study (n=22,872; n_incident_cases_=3,307) (<u>Supplementary Table 10</u>). The two top-performing scores were those derived with LDpred, which comprised 2,324,653 variants (“2021 PRS”) and 1,532,758 variants (“2015 PRS”; <u>Supplementary Tables 11-14</u>). In bootstrapping analyses, the 2021 PRS outperformed the 2015 PRS as evidenced by greater effect estimates (age- and sex-adjusted mean hazard ratio [HR]=1.56 versus 1.49; p-value=3.2×10^−31^) and higher area under the receiver operator characteristic curve (AUC; age- and sex-adjusted mean AUC=0.742 versus 0.736; p-value=6.5×10^−16^) (<u>Supplementary Table 15</u>).

We validated both scores in a held-out subset of the Malmo Diet and Cancer study (n=5,685; n_incident_cases_=815) (<u>Supplementary Table 10</u>). The 2021 PRS was more strongly associated with incident CAD with greater age- and sex-adjusted hazards per 1-SD higher PRS (HR 1.61; 95% CI 1.50-1.72) than the 2015 PRS (HR 1.49 per standard deviation; 95% CI 1.39-1.59), providing improved stratification of participants at higher and lower risk for incident CAD (**Figure 2a**). After adjustment for several established risk factors (total cholesterol, HDL cholesterol, systolic blood pressure, body mass index, type 2 diabetes, current smoking status, and family history of CAD), the 2021 PRS remained strongly associated with incident events (HR 1.54 per SD higher PRS; 95% CI: 1.42-1.66). Examining the extremes of CAD risk, the 2021 PRS yielded a 5.7-fold higher risk between the top and bottom deciles of the PRS, compared to a 3.8-fold higher risk with the 2015 PRS.

**Figure 2.**
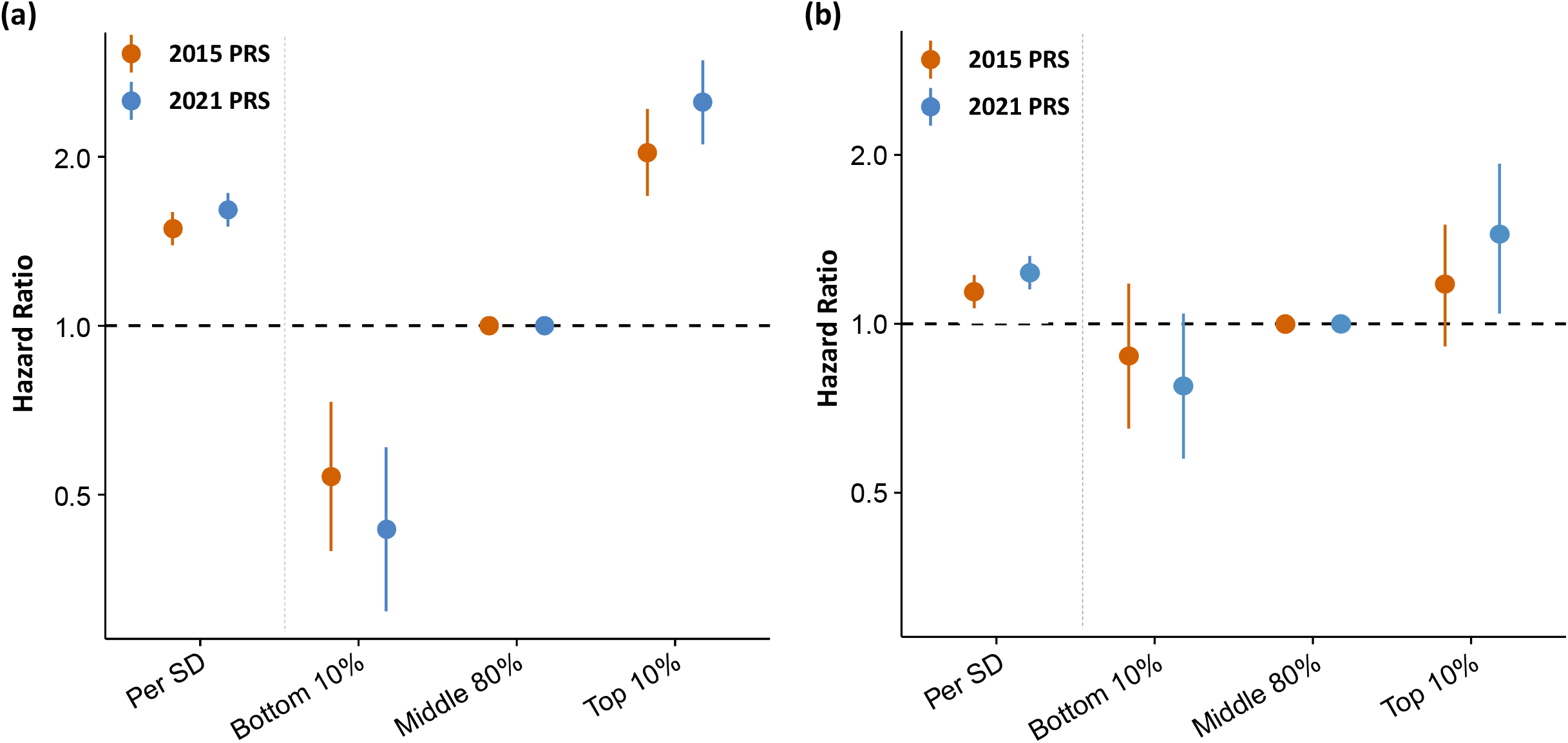
Polygenic prediction of primary and secondary coronary artery disease. Prognostication of (a) incident coronary artery disease and (b) recurrent coronary events by optimal polygenic risk scores derived from the current meta-analysis of ∼180K CAD cases (”2021 PRS” – includes ∼2.3 million variants) or a previously reported GWAS meta-analysis of CAD from 2015 involving ∼60K CAD cases (”2015 PRS” – includes ∼1.5 million variants). 815 incident events were analyzed in the validation subset of the Malmo Diet and Cancer Study and 1,074 recurrent coronary events were analyzed in the FOURIER trial. Cox proportional hazards models were adjusted for age, sex and genetic principal components. Error bars represent 95% confidence intervals of hazard ratio estimates.

To assess the value of the PRS for secondary prevention, we evaluated both PRS for prediction of recurrent coronary events in the placebo arm of the Further Cardiovascular Outcomes Research with PCSK9 Inhibition in Subjects with Elevated Risk (FOURIER; n=7,135; n_incident_cases_=673) clinical trial, a cohort of patients with established atherosclerotic cardiovascular disease.[29] The 2021 PRS demonstrated improved recurrent event prediction (HR 1.20 per SD higher PRS; 95% CI: 1.11-1.29) as compared to the 2015 PRS (HR 1.13 per SD higher PRS; 95% CI: 1.04-1.22), and enhanced stratification of participants at higher and lower risk for secondary events (**Figure 2b**). Examining the extremes of risk, the 2021 PRS yielded a 1.7-fold higher risk of recurrent coronary events between the top and bottom deciles of the PRS versus a 1.4-fold higher risk with the 2015 PRS.

### Prioritizing causal variants, genes and intermediate pathways

We employed several independent approaches to prioritize causal variants, effector genes, relevant tissues of action and related intermediate causal pathways for all 241 genome-wide significant associations. Presence of a protein-altering (i.e. missense or predicted loss-of-function) variant has been shown to be a strong predictor of a causal gene, particularly if the coding variant is not common in the population[14]. At 44 of the 241 genome-wide significant associations, the sentinel variant, or a strong proxy (r^2^ ≥0.8), was a protein-altering variant (<u>Supplementary Table 16</u>). These included well-known low-frequency missense variants in *PCSK9* (p.R46L), *LPL* (p.N291S), and *ANGPTL4* (p.E40K)[16]. Eleven of the 44 missense variants were novel, including a missense variant in *RRBP1* (rs1132274; p.R891Q) that was also the CAD sentinel variant. *RRBP1* encodes ribosome binding protein 1, a widely-expressed protein responsible for protein processing in the membrane of the endoplasmic reticulum. We also identified a missense variant (rs129415; p.G398R) in *SCUBE1* that is strongly correlated with the CAD sentinel variant in European ancestry participants (r^2^ =0.99). *SCUBE1* encodes signal peptide-CUB-EGF domain-containing protein 1, a glycoprotein secreted by activated platelets that protects against thrombosis in mice when inhibited.[30]

### Functionally-informed fine-mapping

Incorporating functional annotations into fine-mapping approaches has been shown to improve identification of likely causal variants at associated loci.[31-33] Using ChromHMM-derived chromatin states from the NIH Roadmap Epigenomics Consortium to functionally annotate the genome, we found greater than 2-fold enrichment for these states in the ten CAD relevant cell/tissue types we tested, consistent with the findings in a previous GWAS meta-analysis (<u>Supplementary Table 17</u>).[7] Of 197 distance-based regions containing genome-wide significant associations, we found 116 (58.9%) regions with significant enrichment in at least one tissue type (<u>Supplementary Table 18</u>). The majority (69; 59.5%) were relatively tissue-specific, showing enrichment in only one or two tissue types, but eight regions showed widespread enrichment in seven or more tissues (**Figure 3a**). Adipose (n=28), liver (n=24) and aorta (n=21) were the tissues that showed the greatest enrichment for the most regions, reflecting their importance in the etiology of CAD (<u>Supplementary Table 18</u>).

**Figure 3.**
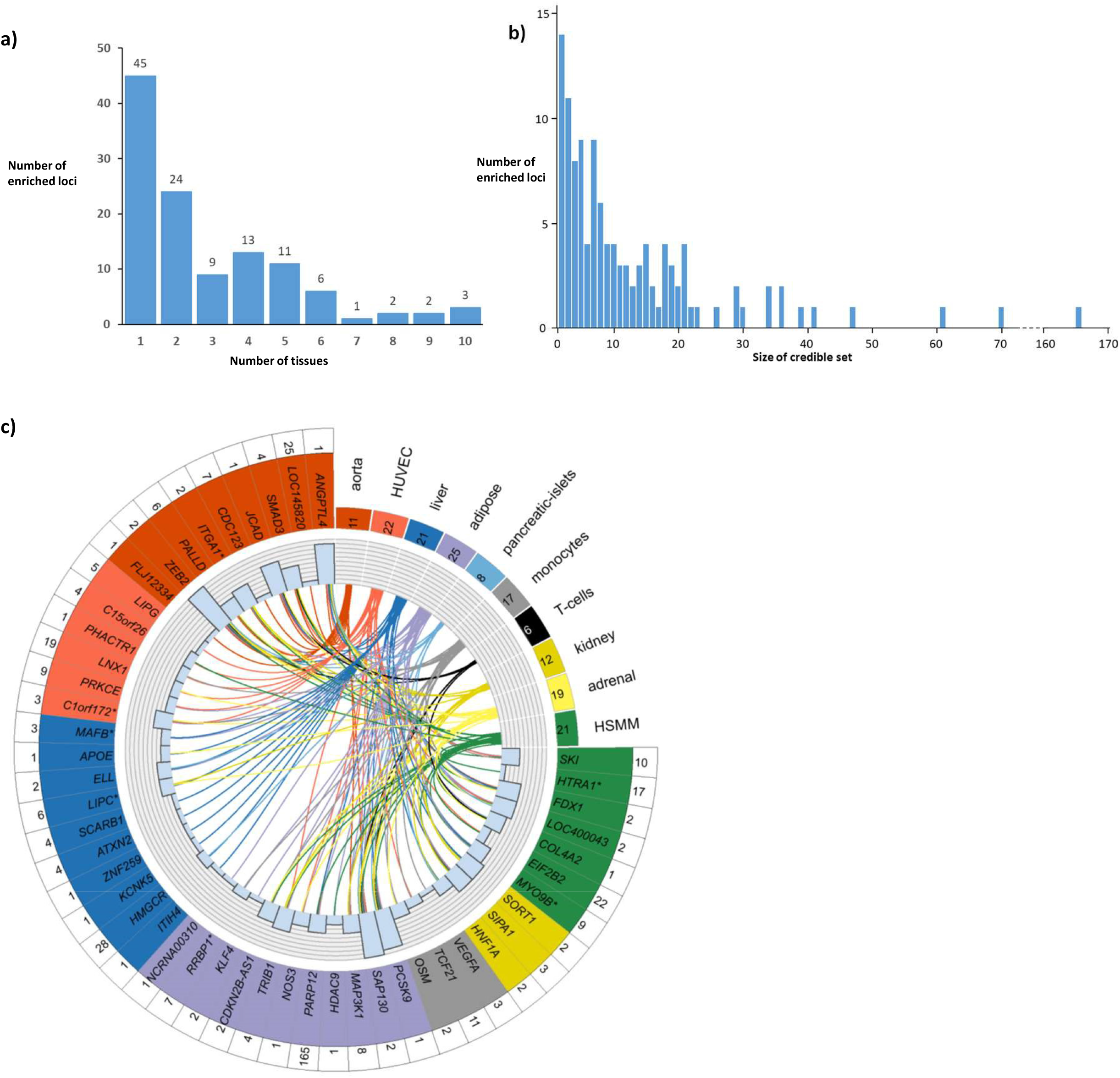
Epigenetic enrichment and functionally-informed fine-mapping of CAD loci. **a) Number of tissues/cell-types the 116 regions were enriched in**. **b) Distribution of 95% credible set sizes for the 116 enriched regions**. **c) Circos plot of epigenetic enrichment for 49 significantly enriched GWAS regions containing a variant with PPA≥0**.**5**. The number of regions each tissue showed enrichment in is displayed in the upper right quadrant. The number of regions that show enrichment with a given tissue/cell-type is displayed in the box next to the tissue/cell-type name. The 49 significantly enriched GWAS regions containing a variant with PPA≥0.5 are colored according to the tissue with the strongest evidence of enrichment for that region. Region names with an asterisk (*) denote those for which all conditionally independent association signals were annotated as being novel. The histogram shows the total number of tissues with enrichment for each region and the links indicate the tissues/cell-types each region was enriched in. The number of 95% credible variants per region is displayed in the outer ring. HSMM = human skeletal muscle myoblasts; HUVEC = human umbilical vein endothelial cells.

We applied a functionally-informed fine-mapping method (FGWAS),[32] which uses the chromatin state enrichment information to reweight GWAS summary statistics and compute variant-specific posterior probabilities of association (PPA). Across the 116 enriched regions we identified 1,456 potential causal variants among the 95% credible sets (<u>Supplementary Table 19</u>). Forty-two enriched regions contained fewer than five 95% credible variants (**Figure 3b**), while 49 regions contained a variant with posterior probability of association (PPA)≥0.5 (**Figure 3c**; <u>Supplementary Table 20</u>), showing that the combination of functional annotation and high statistical power can pinpoint likely causal variants. Indeed, 13 regions were fine-mapped to just a single variant credible set, including missense variants in *PCSK9, ANGPTL4* and *APOE*, plus other well-studied non-coding variants, such as rs9349379 near *PHACTR1/EDN1*,[34] and rs2107595 near *HDAC9/TWIST1*.[35]

At 10 loci, functionally-informed fine-mapping prioritized variants that did not have the strongest statistical association. For example, at the LDL-cholesterol and adiposity-associated *MAFB* locus,[36] the CAD sentinel variant was rs2207132 (OR=1.10, 95%CI=1.07-1.13; p-value=6.7×10^−10^) (<u>Supplementary Table 2</u>; <u>Supplementary Figure 5a</u>). However, a strongly correlated variant (rs1883711; r^2^ =0.92) lies in a region annotated as a likely enhancer in liver and adipose tissue, the two enriched tissues at this locus (<u>Supplementary Figure 5b</u>). Therefore, rs1883711 was strongly upweighted by FGWAS resulting in a PPA of 0.77 compared to 0.13 for rs2207132. We queried CAD-associated variants for *cis*-eQTLs in CAD-relevant tissues from the STARNET and GTEx studies (<u>Online Methods</u>).[37, 38] The eQTL for *MAFB* observed in liver samples from CAD patients in STARNET suggests that the CAD association is mediated by changes in expression of *MAFB* (encoding MAF bZIP transcription factor B) (<u>Supplementary Table 20</u>). MafB expression in macrophages is upregulated by oxidized LDL stimulation,[39] while MafB deficiency in mice has been shown to increase atherosclerosis by inhibiting foam cell apoptosis.[40]

### Polygenic prioritization of candidate causal genes (PoPS)

Combining locus- and similarity-based approaches has been shown to enhance the prioritization of causal genes.[14, 41] However, established similarity-based methods have not leveraged the full polygenic signal to inform gene prioritization. We therefore incorporated a newly developed similarity-based method for gene prioritization, the Polygenic Priority Score (PoPS), which utilizes the full genome-wide association data while excluding a given locus of interest.[15] We applied PoPS to summary-level data from the GWAS meta-analysis using European ancestry individuals from the 1000 Genomes Project as a reference panel.[42] An initial 57,543 features – including data on gene expression, protein-protein interaction networks, and biological pathways – were considered for analysis, of which 21,407 features (37.3%) passed a marginal feature selection step and were input into the final predictive, PoPS model (<u>Online Methods</u>). We computed a PoPS score for all protein-coding genes within a defined 500kb window around each of the 241 genome-wide associations and prioritized the gene with the highest PoPS score in each locus, resulting in 196 prioritized genes. Despite not incorporating locus-specific information, PoPS prioritized many well-established genes implicated in CAD pathogenesis including *LDLR, APOB, PCSK9, SORT1, NOS3, VEGFA*, and *IL6R* (<u>Supplementary Tables 21 & 22</u>).

Next we evaluated groups of features from the final PoPS model to identify those features that were most informative in prioritizing CAD-relevant genes. Hierarchical clustering of the 21,407 features yielded 3,149 clusters, which we ranked by their relative contribution to the PoPS scores of prioritized genes (**Figure 4a**). The highest-ranking cluster contained features indicating homeostatic regulation of blood lipids (<u>Supplementary Table 23</u>). Other top clusters included features related to: function and proliferation of endothelial and smooth muscle cells; the structure and function of the extracellular matrix; and numerous metabolic pathways including those in adipose tissue controlling thermoregulation and turnover of lipids or phospholipids, all well-established pathways and mechanisms in the pathogenesis of CAD[43-45]. In addition, several high-ranking clusters highlighted early developmental processes and signaling pathways involving the cell cycle as less recognized, but important, mediators of CAD risk.

**Figure 4:**
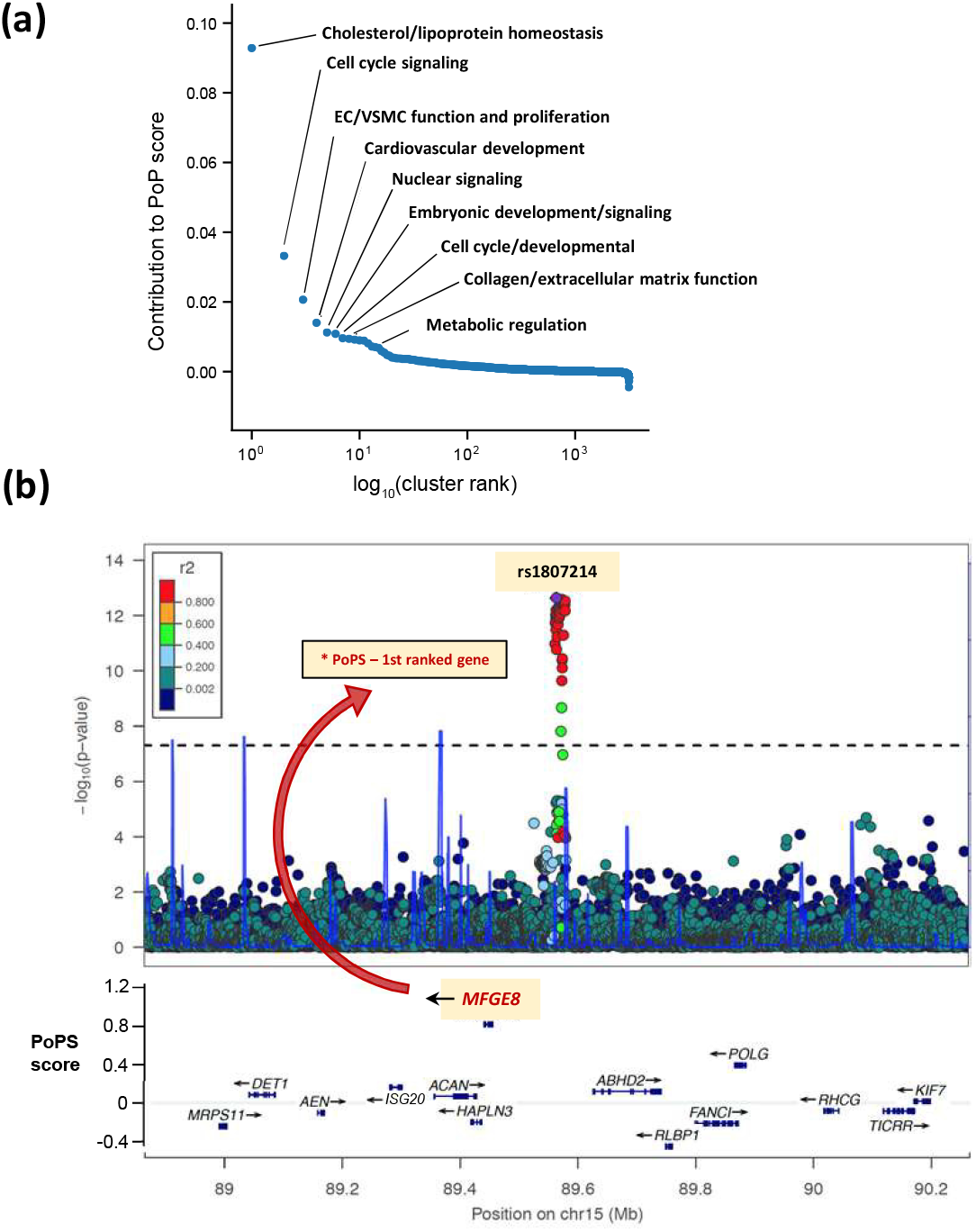
Polygenic priority score (PoPS) informs the identification of causal genes for coronary artery disease (CAD). **(a) Feature clusters contributing to causal gene prioritization**. Rank-order plot of 3,149 feature clusters (arising from 21,407 distinct features) contributing to the prioritization of likely causal genes for CAD by PoPS. Similarity-based cluster labels are provided for several top clusters. **(b) Prioritization of *MFGE8* for rs1807214**. Regional association plot at chromosome 15 demonstrating the prioritization of *MFGE8* as the likely causal gene for rs1807214, which lies in an intergenic region of chromosome 15. Genes in the region are plotted by their chromosomal position (X-axis) and PoPS score (Y-axis).

We then focused on individual loci where the PoPS method informed the prioritization of putative causal genes. For example, rs1807214 was previously reported as genome-wide significant for CAD, but lies in an intergenic region of chromosome 15 at which a causal gene has not been established.[7, 8] Gene expression data from GTEx and STARNET identified *cis*-eQTLs for *ABHD2, MFGE8*, and *HAPLN3* (<u>Supplementary Tables 24 & 25</u>). Prior algorithms combining locus-based approaches have prioritized the nearest gene, *ABHD2*, located 65kb downstream of the sentinel variant.[5, 41] However, PoPS prioritized *MFGE8*, located 108kb upstream of the sentinel, as the most likely causal gene of the ten within 500kb (**Figure 4b**). *MFGE8* encodes lactadherin, an integrin-binding glycoprotein implicated in vascular smooth muscle cell (VSMC) proliferation and invasion, and the secretion of pro-inflammatory molecules.[46, 47] Recently, *in vitro* deletion of this intergenic region by CRISPR/Cas9 was found to increase *MFGE8* expression – with no change to *ABHD2* expression – and *MFGE8* knockdown was shown to reduce coronary artery smooth muscle cell and monocyte (THP-1) proliferation, lending functional support to *MFGE8* as a likely causal mediator of the CAD association in this region.[48]

### Systematic prioritization of putative causal genes

We applied a consensus-based prioritization framework involving eight similarity-based or locus-based predictors to systematically prioritize likely causal genes for all 241 genome-wide associations (<u>Online Methods</u>; **Figure 5a**). Most likely causal genes were selected for each CAD-associated region based on the highest (unweighted) number of the eight predictors. To test this framework, we generated an *a priori* set of 30 “positive control” genes with established causal roles in CAD and assessed the accuracy of each predictor (<u>Supplementary Table 26</u>). 28 of the 30 positive control genes were correctly prioritized as the most likely causal gene based on the highest number of concordant predictors with a median of four concordant predictors per gene (<u>Supplementary Table 27</u>). All predictors demonstrated a high degree of accuracy, including nearest gene (87%), PoPS (80%), eQTL (79%) and mouse knock-outs (95%) (<u>Supplementary Table 27</u>).

**Figure 5.**
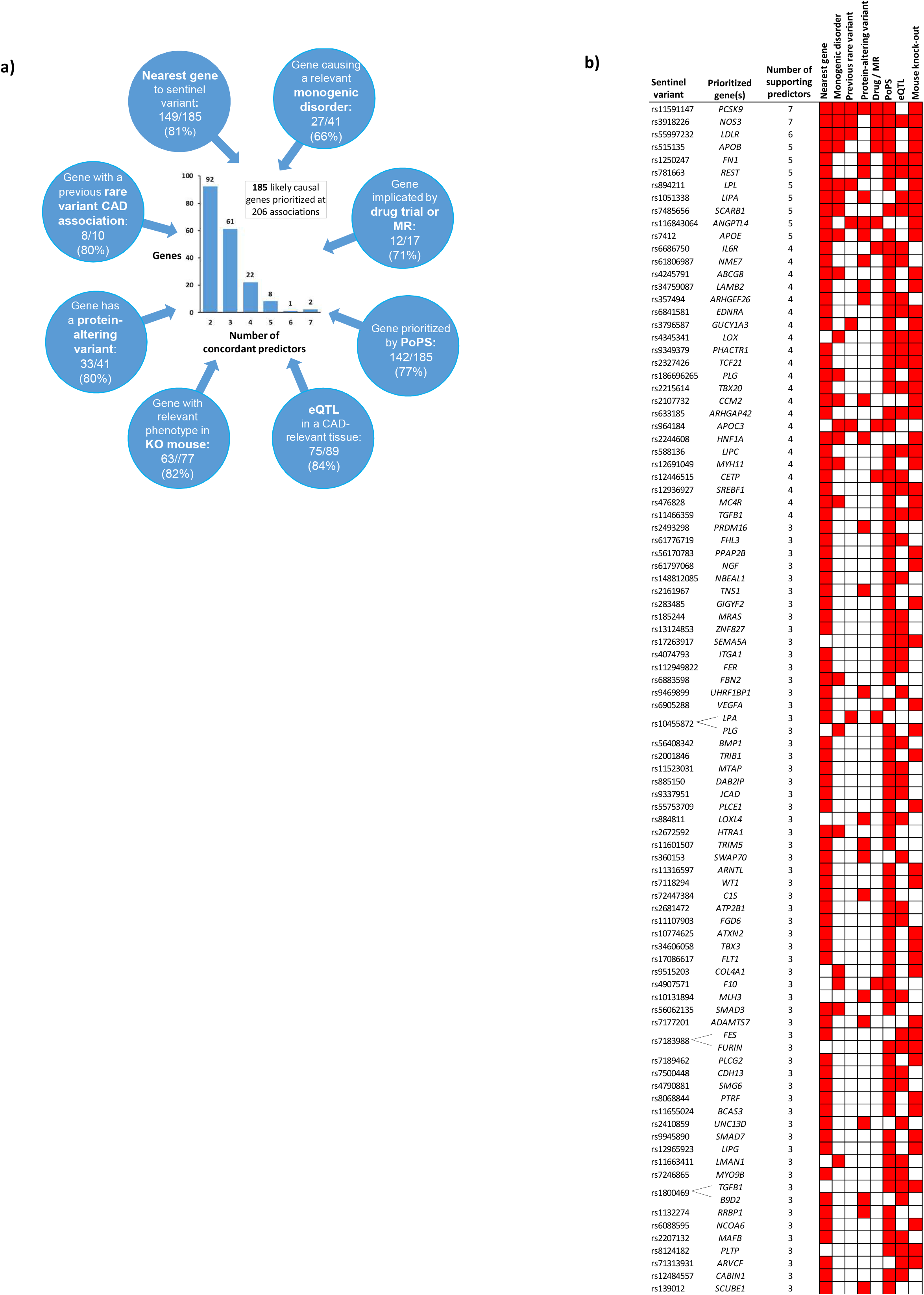
Integrating eight gene-prioritization predictors to identify most likely causal genes. **a) Prioritization of 185 likely causal genes using eight predictors**. Blue circles represent the eight predictors used to prioritize causal genes, which are: 1) A gene in the region harbors a variant that ClinVar classifies as having evidence for being pathogenic for a cardiovascular-relevant monogenic disorder (<u>Supplementary Table 31</u>); 2) A gene in the region has been implicated by an effective drug targeting the protein and/or a positive Mendelian randomization (MR) study suggesting a causal effect of the protein on CAD <u>(Supplementary Table 28</u>); 3) Either of the two top prioritized genes in the region from PoPS (<u>Supplementary Table 21</u>); 4) A gene in the region has an eQTL in a CAD-relevant tissue from GTEx or STARNET for which the lead eSNP is in high LD (r^2^ ≥0.8) with the CAD sentinel variant (<u>Supplementary Tables 24 & 25)</u>; 5) A gene for which a mouse knock-out has a cardiovascular-relevant phenotype (<u>Supplementary Table 32)</u>; 6) A gene in the region harbors a protein-altering variant that is in high LD (r^2^ ≥0.8) with the CAD sentinel variant (<u>Supplementary Table 28</u>); 7) A gene in the region has been shown to have a rare variant association with CAD in a previous whole-exome sequencing (WES) or genotyping study <u>(Supplementary Table 28</u>); 8) The nearest gene to the CAD sentinel variant. Numbers in the blue circles indicate, firstly, the number of genes for which this predictor agreed with the most likely causal gene, secondly, the number of genes for which this predictor provided evidence for at least one gene, and in parentheses, the percentage agreement (i.e. the first number as a percentage of the second). The central histogram shows the number of agreeing predictors that supported the 185 prioritized genes by the number of genes. **b) Predictors for 94 most likely causal genes strongly prioritized by at least three agreeing predictors**. The matrix denotes predictors that supported the mostly likely causal gene (colored red) for each of 94 most likely causal genes with at least three predictors that supported the gene. Genes are ordered by number of agreeing predictors. Lines denote three associations for which two genes were tied for the highest number of agreeing predictors. The sentinel variant for the association with the smallest P-value for CAD is shown for each gene. Full details of the causal gene prioritization evidence for all 241 genome-wide associations are presented in <u>Supplementary Table 28</u>.

We were able to prioritize at least one likely causal gene at 206 (85.5%) of the genome-wide associations based on having at least two concordant predictors, and resulting in the prioritization of 185 distinct genes (<u>Supplementary Table 28</u>). We considered 94 of these genes *strongly* prioritized per the presence of three or more concordant predictors (**Figure 5b**). Overall, for 25 genes, the prioritized gene was not the nearest gene to the sentinel variant, including *APOC3, PLTP* and *LOX. Agreement*, defined as the proportion of times that a predictor prioritized the same gene as the most likely causal gene, was high (but imperfect) across predictors, including nearest gene (149 out of 185; 81%), PoPS (142 out of 185; 77%) and eQTLs (75 out of 89; 84%) (**Figure 5a**). *Concordance*, defined as the proportion of times a pair of predictors both provided evidence for the consensus-based causal gene, was variable (<u>Supplementary Figure 6</u>). For example, nearest gene and presence of a missense variant were typically concordant (30/41, 73%) whereas monogenic genes and eQTL converged on the consensus-based causal gene much less frequently (5/17, 29%).

### Candidate loci with converging variant- and gene-level evidence

Several newly-identified CAD risk loci had strong variant- and gene-level evidence supporting their candidacy for functional interrogation. For example, we identified a CAD-associated region on chromosome 5 that was most strongly enriched in aorta (<u>Supplementary Table 2</u>), and had a 95% credible set of just two variants, with an intronic variant (rs4074793) in *ITGA1* having a PPA of 0.95 (**Figure 6a**,**b**). rs4074793 lies in a region annotated as a likely enhancer in several tissues, and is the lead variant for a strong *cis*-eQTL for *ITGA1* in liver among CAD patients from STARNET (p-value=1.8×10^−73^) (**Figure 6c**). This eQTL was also seen in aorta, subcutaneous fat and mammary artery (**Figure 6d**). No other gene expression signals were seen at this locus, while PoPS also strongly prioritized *ITGA1* as the likely causal gene (<u>Supplementary Table 28</u>). *ITGA1* encodes integrin subunit alpha-1, a widely-expressed protein that forms a heterodimer with integrin beta 1 and acts as a cell surface receptor for extracellular matrix components, such as collagens and laminins. The CAD risk allele (rs4074793-G), or strong proxies, were associated with elevated liver enzymes,[49] C-reactive protein and LDL-cholesterol,[50] highlighting the influence of altered *ITGA1* expression in the liver on lipid pathways as a likely causal pathway to CAD.

**Figure 6.**
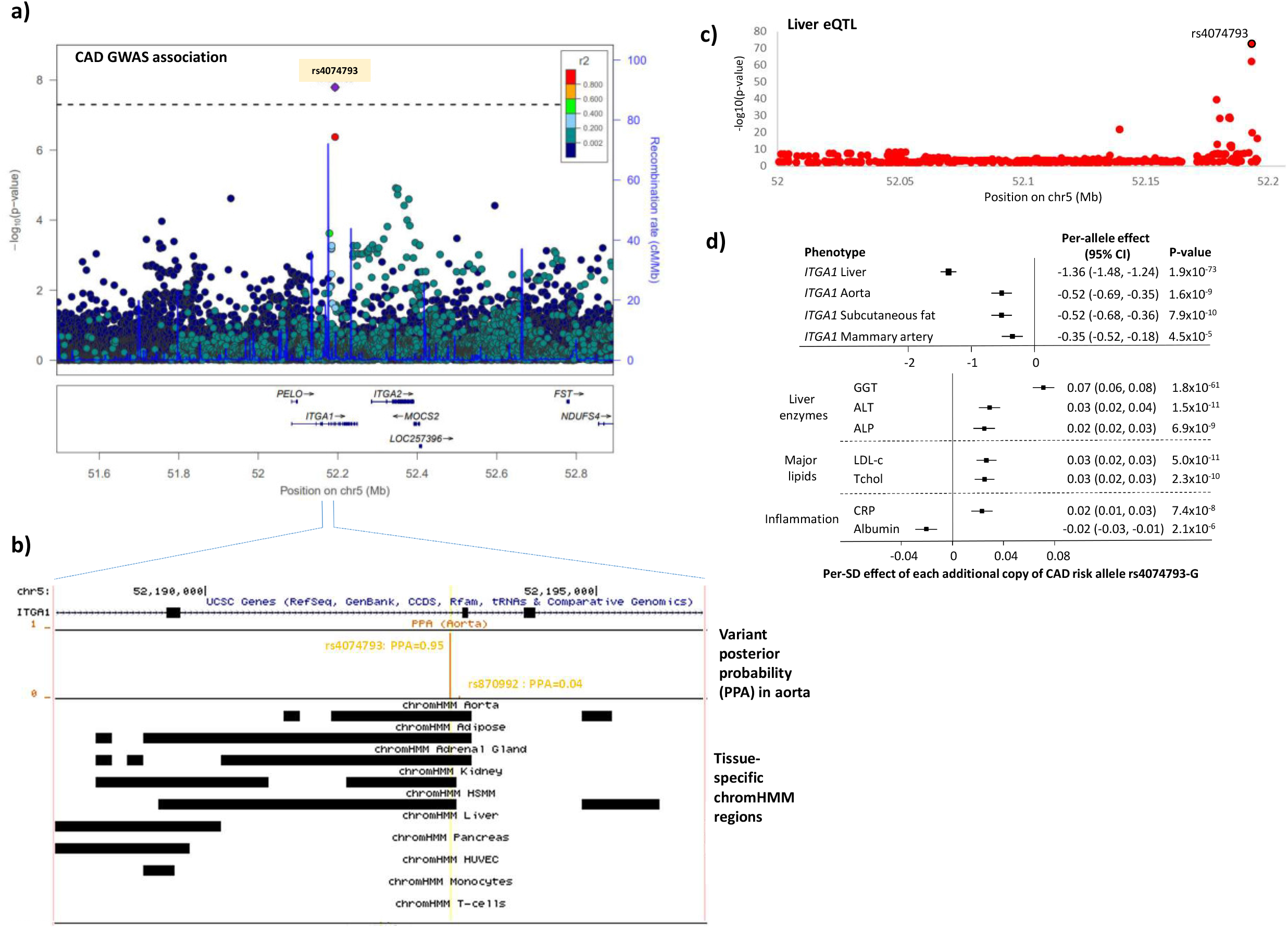
Prioritizing the likely causal variant, gene and pathway at the *ITGA1* locus. **a) Regional association plot from the primary CAD meta-analysis for the *ITGA1* region**. Colored dots represent the position (X-axis) in GRCh37 coordinates and –log10(meta-analysis p-value) (Y axis) of each variant in the region. Dots are shaded to represent the r^2^ with the lead CAD variant (rs4074793), estimated using a random sample of 5,000 European ancestry participants from the UK Biobank. Recombination peaks are plotted in blue based on estimates of recombination from 1000 Genomes European ancestry individuals. **b) Tissue-specific imputed chromHMM states at the two credible set variants in the *ITGA1* region**. The top track shows the position on chromosome 5 (GRCh37) with respect to the *ITGA1* gene. The second track shows as a vertical orange line the posterior probability (Y-axis) for each variant in the region from the FGWAS fine-mapping, identifying rs4074793 (PPA=0.95) as the likely causal variant. The third track indicates as a black box the position of an enhancer state in each of the 10 CAD-relevant tissues, using custom imputed chromHMM states based on epigenomic data from the NIH Roadmap Epigenomics Consortium project. The yellow vertical line indicates the position of the likely causal variant (rs4074793) with respect to the chromHMM states. rs4074793 is annotated to a chromHMM state for all five tissues that show enrichment in the region. HSMM = human skeletal muscle cells; HUVEC = human umbilical vein endothelial cells; PPA = posterior probability of being the causal variant **c) Effect of rs4074973 on *ITGA1* expression in liver in the STARNET study**. The plot shows the position (X-axis) in GRCh37 coordinates and –log10(p-value) (Y axis) of each variant in the region. The likely causal CAD variant rs4074973 is circled in black. **d) Associations of rs4074973 with *ITGA1* expression and phenotypes from a phenome-wide association study**. The per-allele association of rs40747973-G (the CAD risk allele) measured in SD units is plotted for each phenotype. The box indicates the point estimate and the horizontal bars represent the 95% confidence intervals. The top panel shows the association estimates for *ITGA1* expression from the STARNET study. The bottom panel shows associations from UK Biobank (liver enzymes and inflammatory markers) and the literature (lipids: Klarin *et al*., *Nat Genet*, 2018). ALP = alkaline phosphatase; ALT = alanine aminotransferase; CRP = C-reactive protein; GGT = gamma glutamyltransferase; LDL-c = low-density lipoprotein cholesterol; Tchol = total cholesterol.

We identified a novel CAD association near *LIPC* (sentinel variant rs588136, p-value=7.0×10^−10^ ; <u>Supplementary Table 2</u>) where the risk allele was associated with *higher* levels of HDL-cholesterol, opposite the established observational association. *LIPC* encodes hepatic triacylglycerol lipase, a liver-expressed enzyme that catalyzes the hydrolysis of triglycerides and phospholipids in circulating lipoproteins. The region was most strongly enriched for epigenetic annotation in liver, and FGWAS prioritized a 95% credible set comprising 6 variants with rs588316 being the most likely causal variant (PPA=0.50). Variants in the *LIPC* region have been previously associated with circulating HDL-cholesterol levels,[51] but Mendelian randomization studies have reported that HDL-cholesterol is unlikely to play a causal role in CAD risk.[52] Nonetheless, we prioritized *LIPC* as the relevant causal gene per several lines of evidence (<u>Supplementary Table 28</u>): (1) PoPS prioritized *LIPC*, and nine of the 10 strongest features related to lipid and lipoprotein metabolism; (2) *LIPC* is the only gene in the region with a *cis*-eQTL – signals in the liver in both STARNET (p-value=6.0×10^−27^) and GTEx (p-value=1.6×10^−7^); (3) *LIPC* is the only gene in the region with a cardiovascular-relevant phenotype (altered circulating lipid levels) in knock-out mice; (4) the CAD risk allele associated with elevated apolipoprotein-B, LDL-cholesterol and triglycerides in the PheWAS (<u>Supplementary Table 3</u>). The confluence of evidence therefore suggests *LIPC* as the causal gene mediating CAD risk at this region through alterations in liver expression that influence its ability to hydrolyze pro-atherogenic lipids.

Finally, we identified a novel association with CAD at a gene-dense region of chromosome 19 significantly enriched for epigenetic annotations in adipose, liver, monocytes, and skeletal muscle myoblasts (<u>Supplementary Table 2</u>; <u>Supplementary Table 18</u>). FGWAS identified the CAD sentinel variant (rs7246865) as the most likely causal variant (PPA=0.71). PoPS prioritized *MYO9B* as the likely causal gene over 30 other genes within 500kb (<u>Supplementary Table 21</u>). Support was provided by data from GTEx, where the CAD sentinel variant was a *cis*-eQTL (p-value=5.3×10^−8^) for *MYO9B* in tibial artery (<u>Supplementary Table 28</u>). *MYO9B* encodes unconventional myosin-IXb, a myosin protein with Rho-GTPase signaling activity involved in cell migration. Mechanistic *in vitro* and *in vivo* studies have implicated *MYO9B*/RhoA-dependent migration of macrophages in the pathogenesis of abdominal aortic aneurysm, a disease that shares common mechanistic features with CAD.[53] The prioritization of *MYO9B* by PoPS was strongly driven by pathway and PPI-network features pertaining to Rho signaling, proliferation, and chemotaxis (<u>Supplementary Table 21</u>, suggesting a putative causal role for *MYO9B* in CAD pathogenesis, mediated by the proliferation and migration of vascular cell types.

## DISCUSSION

In a genetic discovery analysis involving more than 180,000 cases of CAD and nearly 1 million controls, we identified 241 genome-wide significant associations, including 54 reported here for the first time. We objectively prioritized likely causal variants and effector genes across the 241 associations using functionally-informed fine-mapping, a recently-developed genome-wide gene prioritization method (PoPS), and systematic integration of locus-based and similarity-based predictors, with several tailored specifically to cardiovascular disease.

The large sample size of this study enabled detection of many novel genetic associations with CAD, predominantly weak-effect variants that are common in the population. Our findings suggest that future, larger GWAS - at least those in European ancestry populations - are unlikely to discover many more large-effect common variants (i.e. those with odds ratios greater than 1.05) associated with CAD. In fact, additional associations contributing to the long polygenic tail of CAD risk are likely to arise from the ∼650 predominantly weak effect signals among associations that reached the 1% FDR threshold, which in aggregate explained ∼36% of the heritability of CAD. Notably, we identified 38 additional novel loci - bringing the total number of novel CAD loci reported here to 68 - when we incorporated recently published GWAS results based on only 29,000 CAD cases of East Asian ancestry from Biobank Japan. This observation demonstrates that (future) trans-ethnic genetic analyses should not only identify CAD association signals that differ across ethnicities, but also enhance the yield of overall genetic discovery for CAD.

Consistent with previous studies, we demonstrated that a genome-wide PRS derived from this GWAS strongly predicts both incident and recurrent CAD.[54-57] Notably, the current PRS demonstrated improved ability to discern those at higher and lower risk of CAD as compared to a widely used PRS derived from an earlier GWAS of ∼61,000 CAD cases.[56] While the current PRS provides an improved and powerful tool for genetic risk prediction of CAD in the setting of primary and secondary prevention, our findings suggest that further increases in European-ancestry GWAS sample size may only modestly improve the predictive ability of the CAD PRS. More substantive improvements in polygenic risk prediction may arise from methodological developments, such as approaches that model interactions between variants or incorporate functional information.[58, 59] Moreover, further investigations are required to understand the extent to which genetic discovery analyses that include more non-European ancestry participants will improve the transethnic portability of PRS (and whether this will result in improved prediction across all ancestry groups).[60]

The weak effects of most CAD-associated variants do not preclude their contribution to important etiological insights with therapeutic implications, as the effects of pharmacologically perturbing identified targets are typically much stronger than those of naturally-occurring genetic variants that are common in the population. For example, we uncovered common variant associations of weak effect at *HMGCR* and *NPC1L1*, which encode the targets of HMG-CoA reductase inhibitors (statins) and ezetimibe, respectively, two of the most effective and commonly prescribed medications for the prevention and management of CAD through lowering blood lipid levels. However, the translation of statistical associations into actionable biology and potential therapeutic targets requires elucidation of causal variants, genes and intermediate pathways, which has lagged behind the rapid growth in genetic association discoveries.

Here, we implemented strategies to enhance the identification of putative causal variants and causal genes. By incorporating epigenomic enrichment in disease-relevant tissues - an approach previously shown to improve fine-mapping over broader, disease-agnostic approaches [32] - we prioritized likely causal variants that were not always those with the strongest statistical associations. Using a recently-developed similarity-based tool (PoPS) that exploits the full genome-wide data to identify disease-enriched features, we prioritized likely causal genes for all 241 genome-wide associations. Support for the validity of the genes prioritized by PoPS comes from: the high ranking of features of known relevance to atherosclerosis (e.g. lipid metabolism, extracellular matrix processes) from more than 50,000 tested features; the correct assignment of the most likely causal gene at several well-established lipid and non-lipid CAD loci; selection of the likely-correct causal gene over several other candidates in a region, including those in closer proximity to the sentinel (e.g. *MFGE8*); and corroborating evidence at many loci from orthogonal gene prioritization methods, such as eQTLs in disease-relevant tissues.

As support from multiple, orthogonal lines of evidence increases the likelihood of prioritizing the correct causal gene, we propose an integrative, consensus-based prioritization framework that incorporates eight complementary predictors. By applying this framework to the genome-wide associations with CAD, we provide systematic evidence for the most likely causal gene at over 200 associations. Although distance from the sentinel variant has been shown to be a reasonable predictor of causal genes across many phenotypes,[14, 41] our approach provides added confirmation for many associations. For example, at several newly identified associations, such as those nearest *ITGA1, LIPC* and *MYO9B*, we provide strong support that these proximal genes are most likely causal through both locus-based and similarity-based evidence. However, our framework prioritized a gene that was not the nearest gene for 15% of associations. These included well-known genes such as *APOC3* and *PLTP*, as well as several genes with less well-established, but plausible, roles in CAD, including *MFGE8*. While experimental evidence is required to confirm causal mechanisms, we provide a prioritization framework yielding evidence-based candidates that may be amenable to functional follow-up. Future efforts to improve gene prioritization for CAD may include addition of further disease-specific lines of evidence, such as data from a broader range of relevant cell types (e.g. vascular smooth muscle cells) or high-throughput assays (e.g. genome-wide CRISPR screens).

## METHODS

### Genetic discovery meta-analysis

Details of the ten *de novo* studies, including the source of participants, case and control definitions and basic participant characteristics are presented in <u>Supplementary Table 1</u>. Ethical approval and informed consent were obtained for all participating studies. With the exception of UK Biobank (which used the ThermoFisher UK Biobank Axiom array), studies used Illumina genotyping arrays. Most studies used the Haplotype Reference Consortium v1.1 panel for imputation, but several utilized local whole-genome sequence data for improved imputation. Study-specific sample and variant filters were applied before additive logistic (or logistic mixed) models were run, with CAD status as the outcome and study-specific covariates, including accounting for potential ancestry effects.

We performed an inverse variance weighted meta-analysis on the betas and standard errors using METAL,[61] combining the results from the ten *de novo* studies with previously published summary statistics. To maximize the variant-specific sample size, we used summary statistics from either (a) a previous 1000 Genomes-imputed GWAS meta-analysis of up to 60,801 CAD cases and 123,504 CAD-free controls;[7] (b) a meta-analysis of ∼79,000 variants in up to 88,192 CAD cases and 162,544 controls, predominantly based on the Illumina CardioMetabochip array;[2] or (c) a meta-analysis ∼184,000 variants in up to 42,335 CAD cases and 78,240 controls based on the Illumina Exome array.[10, 16] From each meta-analysis, we dropped variants which were only present in one study or had fewer than 30,000 cases in total from all contributing studies. Where a variant was found in multiple meta-analyses, we kept the result which had the highest total number of ‘effective cases’ across studies (approximated within each study as the variant-specific number of CAD cases multiplied by the imputation quality score). Finally, to avoid false positive associations driven by an extreme result in a single study, we filtered variants with a meta-analysis p-value≤5.0×10^−6^ that did not have a p-value<0.2 in at least two studies for which the direction of effect was consistent with the overall meta-analyses effect estimate. Our final dataset included 20,073,070 variants after filtering.

### Joint association analysis

We performed joint association analysis using GCTA software.[62] This approach fits an approximate multiple regression model using summary-level meta-analysis statistics and LD corrections estimated from a reference panel (here the UKBB sample using European ancestry participants only). We adopted a chromosome-wide stepwise selection procedure to select variants and estimate their joint effects at i) a genome-wide significance level (pJoint≤5.0×10^−8^) in the meta-analyzed variants that reached genome-wide significance (n=18,348), ii) an FDR 1% p-value cut-off (pJoint≤2.52×10^−5^) in the 1% FDR variant list (n=47,622). We identified 241 independent variants at the genome-wide significance threshold and 897 independent variants within the 1% FDR list.

### Identifying previously reported regions and associations

To identify regions of the genome previously reported as having associations with CAD, we first collapsed variants reaching genome-wide significance by clumping variants within 500kb of each other into a single locus. We compared these regions with all variants previously found to be associated with CAD at a genome-wide level of significance (p-value≤5.0×10^−8^) from previous large-scale genetic association studies of CAD. Regions were annotated as ‘known’ if they included a previously reported CAD-associated variant. To assess which of our associations were previously reported or novel, we examined the pairwise correlation between each of our 241 genome-wide significant sentinel variants and any nearby previously reported variants, defining ‘novel’ as having r^2^ <0.2 in UK Biobank European ancestry participants.

### Phenome-wide association study (PheWAS) in UK Biobank

To understand the spectrum of phenotypic consequences of our 241 independent associations with CAD, we conducted a phenome-wide association study in the UK Biobank. For analyses, we selected 53 cardiovascular and non-cardiovascular diseases and 33 continuous traits. A complete list of the phenotypes assessed, details on disease definitions, and relevant sample sizes are provided in <u>Supplementary Tables 29 & 30</u>. We limited analyses to UK Biobank participants of European genetic ancestry as defined by principal components analysis, and excluded one individual in each pair with *KING* coefficient > 0.0884, indicating 2nd degree or closer relatedness (n=393,461). In sensitivity analyses, the PheWAS was repeated after excluding cases of CAD (remaining n=360,255). For disease phenotypes, we performed logistic regression adjusted for age, sex, genotyping array, and the first five principal components. An association with a disease phenotype was deemed significant at a Bonferroni-corrected threshold of p-value<3.9×10^−6^ (53 diseases x 241 genetic variants). Continuous phenotypes were residualized after adjusting for age, sex, genotyping array, and the first five principal components; linear regression was performed on residuals following inverse-normal transformation. For analysis of glycemic traits (hemoglobin A1c and serum glucose), participants with type 1 or type 2 diabetes were excluded. An association with a disease phenotype was deemed significant at a Bonferroni-corrected threshold of P-value<6.3×10^−6^ (33 continuous traits x 241 genetic variants).

### Rare variant analyses

Variant annotation for autosomes was performed using Variant Effect Predictor v96.0 with LOFTEE plugin on version three imputed data and variants with an information score ≥ 0.8.[63, 64] Various gene-based groupings were tested (<u>Supplementary Table 4</u>) and allele frequencies from the entire UK Biobank cohort were used for groupings. Variants (n=64,102) were considered to be in a gene if they fell within the gene coordinates as defined by GENCODE v19. Gene-based association tests were performed in SAIGE-GENE v0.35.8.5 using a white British subset of UK Biobank (28,683 CAD cases and 367,783 controls).[65] Software defaults were used except in step 0 the number of markers for sparse matrix was 2000, and in step 1, the tolerance for preconditioned conjugate gradient to converge was 0.01 and variance ratios were estimated across MAC categories. Two variants were required in each gene for testing. Covariates in the model included the genotyping array, the first five principal components calculated in the white British subset of samples, birth year, and sex. Burden, SKAT, and SKAT-O tests were performed for each gene. As no strong signals were observed except for the *PCSK9* gene, we did not extend our rare variant testing to other studies.

### Sex-specific analysis

We performed a sex-stratified GWAS analysis in UK Biobank following the same phenotype definition and sample exclusions with the main analysis. We used the SAIGE software and adjusted our single-variant association analysis for the first five genetic principal components and the genotyping array, separately for men and women.[66] Based on promising initial results in UK Biobank, we collated sex-stratified GWAS summary statistics from an additional 16 datasets (<u>Supplementary Table 6</u>). All sex-specific summary statistics were checked for quality control (QC) cohort-wise to exclude poorly imputed variants (info<0.4), improbable betas (>|4|) and significant deviations from Hardy-Weinberg Equilibrium (p-value<1.0×10^−9^). Cohort-wise sex-specific q-q plots were generated and inspected and the genomic inflation statistic (λ) was also calculated. Association summary statistics from all 17 studies were combined via inverse-variance weighted meta-analysis in GWAMA.[67, 68] We implemented three different types of meta-analysis: a) a sex-specific meta-analysis, where summary statistics were combined separately for men and women; b) a sex-combined meta-analysis, where effect estimates from men and women were combined assuming no between-sex heterogeneity; and c) a sex-differentiated meta-analysis, where sex-specific estimates were combined while allowing for heterogeneity between men and women. We excluded genetic variants that had a minor allele count < 10 or minor allele frequency < 0.01, were only present in one study, or had a sample size below the median sample size in the sex-combined meta-analysis. To identify significant sex-differentiated genetic variants, we considered variants that had a p-value≤5.0×10^−8^ from the sex-differentiated meta-analysis and a sex-heterogeneity p-value≤0.01. Among the significantly associated genetic variants we then applied a 500kb pruning to identify the sex-differentiated CAD loci.

### False discovery rate (FDR) estimation

The false discovery rate (FDR) following the meta-analysis was assessed using the *‘qvalue’* R package. We generated q-values for all 20.1M variants. The p-value cut-off for a q-value of 1% was 2.52×10^−5^ and there were 47,622 variants reaching that threshold. Joint conditional analysis was performed using GCTA (as described earlier) to identify approximately independent association signals.

### Estimation of heritability explained

Heritability calculations were based on a multifactorial liability-threshold model, implemented in the INDI-V calculator (http://cnsgenomics.com/shiny/INDI-V/), under the assumption of a baseline population risk (K) of 0.0719 and a twin heritability (*H*_L_^*2*^) of 0.4.[69, 70] Single-variant regression estimates from the meta-analysis summary statistics were used to estimate heritability for the sentinel variants at the 241 conditionally independent genome-wide significant associations and the 897 conditionally independent associations reaching the 1% FDR threshold. To account for correlation between variants, multiple regression estimates from the GCTA joint association analysis were also used to estimate heritability for both sets of variants.

### Trans-ethnic comparison

For trans-ethnic comparison we used summary statistics from a recent GWAS of 29,319 CAD cases and 183,134 controls from the Biobank Japan [3]. 199 of the 241 sentinel variants from our primary meta-analysis were also found in the Biobank Japan study; after aligning effect alleles, we compared the beta estimates and minor allele frequencies using Pearson’s correlation coefficient. To investigate the effect of outliers on the between-ancestry correlation of beta estimates, we re-estimated the correlation coefficient after excluding three strong outliers (at *ATXN2, FER*, and *SLC22A1)*. We then performed an inverse variance weighted meta-analysis on the beta estimates and standard errors, incorporating summary results from Biobank Japan and those from all other studies in our primary meta-analysis. After trans-ethnic meta-analysis, we again dropped variants which were only present in one study or had fewer than 30,000 cases in total from all contributing studies, leaving 23,333,163 variants after filtering. We then collapsed variants reaching genome-wide significance (p-value≤5.0×10^−8^) by clumping variants within 500kb into a single locus.

### Derivation and training of polygenic risk scores

Polygenic risk scores (PRS) were derived using one of two methods – pruning and thresholding or the LDpred computational algorithm (LDpred v.1.0), using 503 European ancestry individuals derived from the 1000 Genomes Project study as the linkage disequilibrium reference panel.[42] To evaluate the added utility of our GWAS for the prognostication of CAD risk, we compared two sets of scores using effect estimates from either the current meta-analysis or from our previous 1000 Genomes-imputed GWAS of CAD involving ∼60,000 cases.[7] For each derivation method and summary statistic, we constructed a range of scores of varying sizes drawing from common genetic variants that overlapped between the current meta-analysis, the earlier 1000 Genomes-imputed CAD GWAS, and our training/validation datasets from the Malmö Diet and Cancer (MDC) Study.

Pruning and thresholding-based scores were created using a combination of p-value (1, 0.5, 0.05,5×10^−3^, 5×10^−4^, 5×10^−5^, 5×10^−6^, 5×10^−7^, 5×10^−8^) and r^2^ (0.2, 0.4, 0.6, 0.8, 0.99) thresholds, yielding 45 distinct PRS for each of the two GWAS summary statistics utilized (90 total pruning and thresholding-based scores). LDpred-based scores were constructed incorporating all available SNPs, HapMap3 SNPs (gs://gnomad-public/resources/grch37/hapmap/hapmap_3.3.b37.vcf.bgz), or a soft LD-clumping approach using combinations of p-value (0.05, 0.5, 1) and r^2^ (0.2, 0.4, 0.6, 0.8, 0.99) thresholds (17 total sets of input variants). Additionally, we employed a tuning parameter (ρ) for LDpred, which represents the fraction of causal variants, and tested all LDpred-based scores across a range of ρ parameters (1, 0.3, 0.1, 0.03, 0.01, 0.003, 0.001 and an infinitesimal model), yielding 136 distinct PRS per summary statistic utilized (272 total LDpred-based PRS).

PRS were computed using variants with high-quality imputation results available in MDC, defined by information score (INFO) > 0.3. For each participant, the raw PRS was generated by multiplying the genotype dosage for each risk-increasing allele by its respective weight and then summing across all variants in the score using PLINK2 software. To permit adjustment for genetic ancestry, principal components of ancestry were computed using the EIGENSOFT software package. The calculated raw PRS was ancestry-adjusted by taking the residual of a linear regression model that predicted PRS using the first ten principal components.

We trained all pruning and thresholding and LDpred PRS (362 total scores) in a subset of the Malmö Diet and Cancer Study (n=22,872; n_incident_cases_=3,307). Cox proportional hazard models were used to assess the time-to-event relationship between each PRS and incident CAD with or without adjustment for age and sex. Bootstrapping analysis was performed (100 iterations) and the mean hazard ratio (HR) and mean area under the receiver operator characteristic curve (AUC; as calculated by Harrell’s C-statistic) were reported as performance metrics to rank scores within each of four categories as classified by the PRS derivation method (pruning and thresholding; LDpred) and effect estimates utilized (2015 CAD GWAS; Current meta-analysis). Metrics for the top-performing PRS in each category were compared by Wilcoxon rank-sum test based on results of bootstrapping analyses.

### Primary event prediction analyses in the Malmo Diet and Cancer study (MDC)

The Malmö Diet and Cancer Study is a prospective, population-based cohort that enrolled 30,447 participants between 1991 and 1996 ranging in age from 44 to 73 years. Baseline information on lifestyle and clinical factors was collected using a detailed questionnaire as previously described.[71] From the total study population, 28,556 participants (94%) who had genetic data available and were free of CAD at time of enrollment were analyzed. A subset of 5685 randomly selected participants, that comprised the Malmö Diet and Cancer Cardiovascular Cohort, had blood cholesterol concentrations recorded. Incident cases of CAD had either fatal or nonfatal myocardial infarction, coronary artery bypass graft surgery, percutaneous coronary angioplasty or death due to CAD. Incident event adjudication was available through December 31, 2016. Genotyping was performed using the Illumina GSA v1 genotyping array. Of 29,304 samples which underwent genotyping and were free from CAD at baseline, 28 556 (97%) were retained after quality control procedures that removed low-quality samples (discordance between reported and genetically inferred sex, low call rate (<90%), and sample duplicates). With respect to genetic variants, quality control was performed with removal of those not in Hardy-Weinberg equilibrium (p-value<1×10^−15^). Imputation was then performed using the Haplotype Reference Consortium reference panel.

Cox proportional hazard models were used to assess the time-to-event relationship between each PRS and incident CAD events; baseline models were adjusted for age and sex only, and then subsequently for established risk factors for CAD (total cholesterol, HDL cholesterol, systolic blood pressure, body mass index, type 2 diabetes, current smoking status, and family history of CAD). Harrell C-statistics were estimated using Cox proportional hazard analysis over a 21-year follow-up period to assess the discrimination of the PRS.

### The FOURIER trial (and genetic subset)

The FOURIER trial was a multinational, randomized, double-blind, placebo-controlled trial of the efficacy of evolocumab in patients with clinically evident atherosclerotic cardiovascular disease.[29] The key inclusion criteria for the trial were age between 40 and 85 years, LDL cholesterol of 70 mg/dl or greater or non-HDL-C of 100 mg/dl or greater, and a history of either myocardial infarction, non-hemorrhagic stroke, or symptomatic peripheral artery disease. The genetic sub-study included all participants in FOURIER who provided consent for genetic analyses at enrollment into the trial and had genotyped data that passed quality control (QC), and were of European ancestry. The final genetic cohort comprised of 14,298 unrelated European-ancestry participants, of whom 7,135 were in the placebo arm of the trial. There were no clinically important differences between the overall trial participants and the participants in the genetic subset.

### Secondary event prediction analyses in the FOURIER trial

The two optimal PRS (“2021 PRS” and “2015 PRS”) were calculated using the genotype dosage for each allele, multiplied by its weight, and then summed across all variants. Patients received a raw score standardized per 1-SD (continuous), as well as a percentile score relative to the total cohort. All scoring was performed using PLINK v2.0 (www.cog-genomics.org/plink/2.0/).[72] Model goodness-of-fit was evaluated using the concordance statistic and the Akaike’s Information Criterion (AIC). R version 3.6.1 was used for statistical analyses.

The clinical outcome of interest was recurrent major coronary events, defined as myocardial infarction, coronary revascularization or death from CAD (n_incident_cases_=673). Participants in the genetic cohort were followed for a median of 2.3 years. All endpoints were formally adjudicated by a blinded clinical events committee during the trial. A Cox model was used to determine the hazard ratio per 1 standard deviation higher level of the polygenic risk score and for the extreme deciles compared to the middle 80%. Analyses were adjusted for age, sex, and ancestry (using principal components 1-5).

### Identifying protein-altering variants

To identify protein-altering variants among our genome-wide significant associations, we took the 241 sentinel variants and their LD proxies at r^2^ ≥0.8 as estimated in the European ancestry subset of UK Biobank, and annotated them using the Ensembl Variant Effect Predictor (VEP).[64] We selected for each sentinel variant any proxies identified as having a ‘high’ (i.e. stop-gain and stop-loss, frameshift indel, donor and acceptor splice-site and initiator codon variants) or ‘moderate’ (i.e. missense, in-frame indel, splice region) consequence and recorded the gene that the variant disrupts.

### Functional GWAS analysis

To fine-map loci and identify credible functional variants, we applied the FGWAS software.[32] The software integrates GWAS summary statistics with epigenetic data and we used the ChromHMM-derived states from the NIH Roadmap Epigenomics Consortium on a selection of ten CAD relevant cell/tissue types (adipose nuclei, aorta, human skeletal muscle myoblasts [HSMM], liver, human umbilical vein endothelial cells [HUVEC], kidney, adrenal gland, pancreatic islets, primary monocytes and T-cells from peripheral blood).[73, 74] In order to maximize our search space to find functional elements we prepared a custom state by merging likely functional ChromHMM states (enhancers, transcription start sites [TSS], repressed polycomb, transcription at 5’ and 3’ of gene) for each genomic position. We reweighted the GWAS by running a null model and then a model containing the custom annotation for each of the ten tissues. The regions of the genome that showed strong enrichment (>3SD increment in Bayes Factor [BF]) and had a genome-wide significant CAD-associated variant (p-value<5.0×10^−8^) were selected. For each region, we identified the tissue that showed maximum increment in BF and then constructed a 95% credible functional set of variants based on the ranked posterior probability of association (PPA) for each variant within a region.

### Expression QTL analysis in CAD-relevant tissues

To examine whether the CAD associations were driven by changes in gene expression in CAD-relevant tissues and cell types, we interrogated the Stockholm-Tartu Atherosclerosis Reverse Network Engineering Task (STARNET) eQTL study and the Genotype-Tissue Expression (GTEx) study.[37, 38] For STARNET we used *cis*-eQTL associations from seven tissues (atherosclerotic aortic root [AOR], atherosclerotic-lesion-free internal mammary artery [MAM], blood [BLD], liver [LIV], subcutaneous fat [SF], skeletal muscle [SKLM], and visceral abdominal fat [VAF]) taken from 600 CAD patients as previously described. We cross-referenced the sentinel CAD variants and their proxies (r^2^ ≥0.8) with STARNET eQTLs reaching a 5% FDR for all tissues. To ensure the CAD association and eQTL are likely to be driven by the same causal variant, we retained only those eQTLs where the CAD-associated variant and the lead eQTL variant had r^2^ ≥0.8 among European ancestry participants from UK Biobank. For GTEx we followed the same procedure using the v7 data release (https://www.gtexportal.org/home/datasets) and restricted to *cis-*eQTLs reaching a 5% FDR from eight tissues (adipose [subcutaneous, visceral omentum], adrenal gland, artery [aorta, coronary, tibial], liver and whole blood).

### Polygenic prioritization of candidate causal genes (PoPS)

We implemented PoPS, a gene prioritization method designed to leverage the full genome-wide signal to nominate causal genes independent of methods utilizing GWAS data proximal to the gene.[15] PoPS leverages polygenic enrichments of gene features including cell-type specific gene expression, curated biological pathways, and protein-protein interaction networks to compute a polygenic priority score (POPS) and a p-value for each gene without using any genetic association data on the chromosome containing the gene. Specifically, PoPS was used to train a linear model to predict gene-level association scores from gene features. First, PoPS applied MAGMA to GWAS summary statistics using the 1000 Genomes Project reference panel,[42] and computed gene p-values that are derived from the mean chi-square statistic of SNPs within the gene body. The gene p-values were converted to z-scores z_g_=F^-1^ (1 – p_g_), where F^-1^ was the probit function. This yielded a roughly normally distributed variable that reflects the strength of the association each gene has to the phenotype, which PoPS used as the model target. In total, 57,543 features were considered for analysis, including data on gene expression, protein-protein interaction networks, and biological pathways. After marginal feature selection, PoPS used leave one chromosome out (LOCO), generalized least squares, with l2 regularization to learn linear coefficients for the gene features. Finally, using LOCO prediction, PoPS computed a polygenic priority score for each gene.

### Variants responsible for cardiovascular-relevant monogenic disorders

To identify genes harboring pathogenic variants responsible for cardiovascular-relevant monogenic disorders, we searched the NCBI’s ClinVar database (https://www.ncbi.nlm.nih.gov/clinvar/) on 26th June 2020. Variants were pruned to those: within ±500kb of our CAD sentinel variants; categorized as ‘pathogenic’ or ‘likely pathogenic’; with a listed phenotype; and with either (a) details of the evidence for pathogenicity, (b) expert review of the gene, or (c) a gene that appears in practice guidelines. We then filtered variants that were annotated with a manually curated set of cardiovascular-relevant phenotype terms, including those related to cardiovascular diseases (CAD, cardiovascular disease), CAD risk factors (lipids, metabolism, blood pressure, obesity, platelets), bleeding disorders and relevant cardiac, vasculature or neurological abnormalities (<u>Supplementary Table 31</u>). Where a variant was annotated with multiple genes, both genes were considered as potentially pathogenic.

### Phenotyping knock-out mice

Human gene symbols were mapped to gene identifiers (HGNC) and mouse ortholog genes were obtained using Ensembl (www.ensembl.org). Phenotype data for single-gene knock-out models were obtained from the International Mouse Phenotyping Consortium, data release 10.1 (www.mousephenotype.org), and from the Mouse Genome Informatics database, data from July 2019 (www.informatics.jax.org). For each mouse model, reported phenotypes were grouped using the mammalian phenotype ontology hierarchy into broad categories relevant to CAD: cardiovascular physiology (MP:0001544), cardiovascular morphology (MP:0002127), growth and body weight (MP:0001259), lipid homeostasis (MP:0002118), cholesterol homeostasis (MP:0005278), and lung morphology (MP:0001175). This resulted in mapping from genes to phenotypes in animals (<u>Supplementary Table 32</u>).

### Rare variant associations, Mendelian randomization and drug evidence

To inform prioritization of causal genes within 1Mb regions around our genome-wide associations, we reviewed the literature for three sources of evidence: (1) rare coding variants previously associated with CAD, either individually or in aggregate gene-based tests, through whole-exome sequencing or exome array studies; (2) Mendelian randomization studies of gene expression, protein levels or proximal phenotypes that implicate specific genes as causal effector genes for CAD; (3) drugs proven to be effective for cardiovascular-relevant indications and that target specific proteins encoded by genes.

### Systematic integration of gene prioritization evidence

To systematically prioritize likely causal genes for all 241 genome-wide associations, we integrated eight of the aforementioned similarity-based or locus-based predictors of causal genes: (1) the top two prioritized genes from PoPS; (2) genes with eQTLs in CAD-relevant tissues from STARNET or GTEx; (3) genes containing protein-altering variants that are in strong LD (r^2^ ≥0.8) with the CAD sentinel variant; (4) genes harboring variants responsible for monogenic disorders of cardiovascular relevance according to ClinVar; (5) genes containing rare coding variants that have been associated with CAD risk in previous whole-exome sequencing or array-based studies; (6) genes encoding proteins of causal relevance to CAD per Mendelian randomization studies, or that are targets for established cardiovascular drugs; (7) genes that display cardiovascular-relevant phenotypes in knock-out mice from the International Mouse Phenotyping Consortium or Mouse Genome Informatics database; and (8) the nearest gene to the CAD sentinel variant (**Figure 5a**). We prioritized the most likely “causal gene” for each association using a consensus-based approach, selecting the gene with the highest, unweighted sum of evidence across all eight predictors.

We tested our approach by evaluating whether 30 (“positive-control”) genes with established relevance to CAD were prioritized as the most likely causal genes within their respective genomic regions. In addition, we defined two measures to summarize the relative contributions of individual predictors and pairs of predictors to the consensus-based approach. Specifically, we defined “*Agreement*” as the proportion of times that an individual predictor prioritized the same gene that was nominated as the most likely causal gene by the consensus-based framework. “*Concordance*” was defined as the proportion of times a pair of predictors both converged on the gene that was nominated as the most likely causal gene by the consensus of the eight predictors.

## Data Availability

Summary statistics will be made available upon publication through the CARDIoGRAMplusC4D website (http://www.cardiogramplusc4d.org/) and the NHGRI-EBI GWAS Catalog (https://www.ebi.ac.uk/gwas/) and polygenic risk score weights will be deposited in the Polygenic Score (PGS) Catalog (https://www.pgscatalog.org/). Interactive searchable Manhattan plots and a locus-specific epigenome annotation browser for functionally enriched loci are available at: https://procardis.shinyapps.io/cadgen/. Regional association plots for all 241 genome-wide significant associations are available for download from https://drive.google.com/file/d/1AULeR5zAQJIdR6uNHidJ6xs5AxOe6i5M/view?usp=sharing. An interactive searchable browser detailing the locus-specific evidence prioritizing causal variants, genes and pathways is available at the Common Metabolic Diseases Knowledge Portal (beta version available at: https://hugeamp.org/method.html?trait=cad&dataset=cardiogram).

https://procardis.shinyapps.io/cadgen/

https://hugeamp.org/method.html?trait=cad&dataset=cardiogram

## DATA AVAILABILITY

Summary statistics will be made available upon publication through the CARDIoGRAMplusC4D website (http://www.cardiogramplusc4d.org/) and the NHGRI-EBI GWAS Catalog (https://www.ebi.ac.uk/gwas/) and polygenic risk score weights will be deposited in the Polygenic Score (PGS) Catalog (https://www.pgscatalog.org/). Interactive searchable Manhattan plots and a locus-specific epigenome annotation browser for functionally enriched loci are available at: https://procardis.shinyapps.io/cadgen/. Regional association plots for all 241 genome-wide significant associations are available for download from https://drive.google.com/file/d/1AULeR5zAQJIdR6uNHidJ6xs5AxOe6i5M/view?usp=sharing. An interactive searchable browser detailing the locus-specific evidence prioritizing causal variants, genes and pathways is available at the Common Metabolic Disease’s Knowledge Portal (beta version available at: https://hugeamp.org/method.html?trait=cad&dataset=cardiogram).

## CODE AVAILABILITY

Code used in this project is available on reasonable request to the corresponding authors.

## ACKNOWLEDGEMENTS

T.K. is supported by the Corona-Foundation (Junior Research Group Translational Cardiovascular Genomics) and the German Research Foundation (DFG) as part of the Sonderforschungsbereich SFB 1123 (B02). J.D. is a British Heart Foundation Professor, European Research Council Senior Investigator, and National Institute for Health Research (NIHR) Senior Investigator. J.C.H. acknowledges personal funding from the British Heart Foundation (FS/14/55/30806) and is a member of the Oxford BHF Centre of Research Excellence (RE/13/1/30181). R.C. has received funding from the British Heart Foundation and British Heart Foundation Centre of Research Excellence. O.G. has received funding from the British Heart Foundation (BHF) (FS/14/66/3129). P.S.dV was supported by American Heart Association grant number 18CDA34110116 and National Heart, Lung, and Blood Institute grant R01HL146860. The Atherosclerosis Risk in Communities study has been funded in whole or in part with Federal funds from the National Heart, Lung, and Blood Institute, National Institutes of Health, Department of Health and Human Services (contract numbers HHSN268201700001I, HHSN268201700002I, HHSN268201700003I, HHSN268201700004I and HHSN268201700005I), R01HL087641, R01HL059367 and R01HL086694; National Human Genome Research Institute contract U01HG004402; and National Institutes of Health contract HHSN268200625226C. The authors thank the staff and participants of the ARIC study for their important contributions. Infrastructure was partly supported by Grant Number UL1RR025005, a component of the National Institutes of Health and NIH Roadmap for Medical Research. The Trøndelag Health Study (The HUNT Study) is a collaboration between HUNT Research Centre (Faculty of Medicine and Health Sciences, NTNU, Norwegian University of Science and Technology), Trøndelag County Council, Central Norway Regional Health Authority, and the Norwegian Institute of Public Health. The K.G. Jebsen Center for Genetic Epidemiology is financed by Stiftelsen Kristian Gerhard Jebsen; Faculty of Medicine and Health Sciences, NTNU, Norwegian University of Science and Technology; and Central Norway Regional Health Authority. Whole genome sequencing for the HUNT study was funded by HL109946. The GerMIFs gratefully acknowledge the support of the Bavarian State Ministry of Health and Care, furthermore founded this work within its framework of DigiMed Bayern (grant No: DMB-1805-0001), the German Federal Ministry of Education and Research (BMBF) within the framework of ERA-NET on Cardiovascular Disease (Druggable-MI-genes: 01KL1802), within the scheme of target validation (BlockCAD: 16GW0198K), within the framework of the e:Med research and funding concept (AbCD-Net: 01ZX1706C), the British Heart Foundation (BHF)/German Centre of Cardiovascular Research (DZHK)-collaboration (VIAgenomics) and the German Research Foundation (DFG) as part of the Sonderforschungsbereich SFB 1123 (B02) and the Sonderforschungsbereich SFB TRR 267 (B05). This work was supported by the British Heart Foundation (BHF) grant RG/14/5/30893 (P.D.) and forms part of the research themes contributing to the translational research portfolios of the Barts Biomedical Research Centre funded by the UK National Institute for Health Research (NIHR). I.S. is supported by a Precision Health Scholars Award from the University of Michigan Medical School. This work was supported by the European Commission (HEALTH-F2–2013-601456) and the TriPartite Immunometabolism Consortium [TrIC]-NovoNordisk Foundation (NNF15CC0018486), VIAgenomics (SP/19/2/344612), the British Heart Foundation, a Wellcome Trust core award (M.F., H.W., 203141/Z/16/Z) and support from the NIHR Oxford Biomedical Research Centre. M.F. and H.W. are members of the Oxford BHF Centre of Research Excellence (RE/13/1/30181). The views expressed are those of the authors and not necessarily those of the NHS, the NIHR or the Department of Health. C.P.N. and T.R.W are funded by the British Heart Foundation. C.J.W. is funded by NIH grant R35-HL135824. B.N.W is supported by the National Science Foundation Graduate Research Program (DGE 1256260). This research was supported by BHF (SP/13/2/30111) and conducted using the UK Biobank Resource (application number 9922). O.M. was funded by the Swedish Heart- and Lung Foundation, the Swedish Research Council, the European Research Council ERC-AdG-2019-885003 and Lund University Infrastructure grant “Malmö population-based cohorts” (STYR 2019/2046). T.R.W. is funded by the British Heart Foundation. I.K., S.Ko., and K.It. are funded by the Japan Agency for Medical Research and Development, AMED, under Grant Numbers JP16ek0109070h0003, JP18kk0205008h0003, JP18kk0205001s0703, JP20km0405209, and JP20ek0109487. The BioBank Japan is supported by AMED under Grant Number JP20km0605001. J.L.M.B. acknowledges research support from NIH R01HL125863, American Heart Association (A14SFRN20840000), the Swedish Research Council (2018-02529) and Heart Lung Foundation (20170265) and the Foundation Leducq (PlaqueOmics: Novel Roles of Smooth Muscle and Other Matrix Producing Cells in Atherosclerotic Plaque Stability and Rupture, 18CVD02. A.V.K. has been funded by 1K08HG010155 from the National Human Genome Research Institute. K.G.A. has received support from the American Heart Association Institute for Precision Cardiovascular Medicine (17IFUNP3384001) and a KL2/Catalyst Medical Research Investigator Training (CMeRIT) award from the Harvard Catalyst (KL2 TR002542). EPIC-CVD was funded by the European Research Council (268834) and the European Commission Framework Programme 7 (HEALTH-F2-2012-279233). The coordinating centre was supported by core funding from the: UK Medical Research Council (G0800270; MR/L003120/1), British Heart Foundation (SP/09/002; RG/13/13/30194; RG/18/13/33946) and NIHR Cambridge Biomedical Research Centre (BRC-1215-20014). The views expressed are those of the author(s) and not necessarily those of the NIHR or the Department of Health and Social Care.

## CONFLICTS OF INTEREST

All deCODE affiliated authors are employees of deCODE/Amgen Inc. The TIMI Study Group has received institutional research grant support through Brigham and Women’s from Abbott, Amgen, Aralez, AstraZeneca, Bayer HealthCare Pharmaceuticals, Inc., BRAHMS, Daiichi-Sankyo, Eisai, GlaxoSmithKline, Intarcia, Janssen, MedImmune, Merck, Novartis, Pfizer, Poxel, Quark Pharmaceuticals, Roche, Takeda, The Medicines Company, and Zora Biosciences. R.C., J.C.H., M.I., F.M. work at the Clinical Trial Service Unit and Epidemiological Studies Unit, Nuffield Department of Population Health, which receives research grants from industry that are governed by University of Oxford contracts that protect its independence, and has a staff policy of not taking personal payments from industry; further details can be found at https://www.ndph.ox.ac.uk/files/about/ndph-independence-of-research-policy-jun-20.pdf. A.S.B reports grants outside of this work from AstraZeneca, Bayer, Biogen, BioMarin, Bioverativ, Merck, Novartis and Sanofi. A.B. and L.A.L. are employees of Regeneron Pharmaceuticals and the spouse of C.J.W. works at Regeneron Pharmaceuticals. J.L.M.B. and A.R. are members of the board of directors, founders and shareholders of Clinical Gene Networks AB that has an invested interest in STARNET. J.C.U. has received compensation for consulting from Goldfinch Bio and is an employee of Patch Biosciences. O.G. became a full-time employee of UCB while this manuscript was being drafted.

All other co-authors report no conflicts of interest.

## AUTHOR CONTRIBUTIONS

<u>De novo GWAS</u>: K.G.A., T.J., B.N.W., W.Z., C.R., I.S., L.M.V., B.O.A., D.O.A., A.B., J.D., G.D., P.D., P.T.E., J.E., O.G., P.G., D.F.G., U.G., S.M.I.H., A.H., G.H., H.H., K.H., A.K., S.Kat., T.K., A.K., L.L., N.A.M., T.M., S.M., L.M.V., M.M., J.B.N., M.M.N., S.P., L.S.R., T.R., C.R., C.T.R., M.S.S., H.S., K.S., I.S., G.Thorg., G.Thorl., U.T., M.v.S., and C.J.W.

<u>Discovery meta-analysis</u>: K.G.A., T.J. and A.S.B.

<u>Conditional analysis, FDR analysis, heritability estimation</u>: S.Kan.

<u>Rare variant analysis</u>: B.N.W., W.Z., I.S., and C.J.W.

<u>Sex analysis</u>: S.Kan., P.D., K.G.A., B.O.A., E.B., M.R.B., B.B., A.S.B., R.C., J.D., P.S.deV., J.E., M.F., A.G., C.G., U.G., S.M.I.H., G.H., J.C.W., K.H., M.I., T.J., A.K., S.Kat., T.K., A.K., L.L., T.M., A.C.M., S.M., L.M.V., M.M., F.M., J.B.N., M.M.N., S.P., T.R., H.S., I.S., M.v.S., H.W., C.J.W., B.N.W., P.Z., and W.Z. <u>PheWAS</u>: K.G.A.

<u>Trans-ethnic analysis</u>: K.G.A., T.J., K.Is., K.It., Y.K., I.K., S.Ko., and A.S.B.

<u>Association of polygenic risk scores</u>: K.G.A., M.W., G.H., C.R., N.A.M., F.K.K., L.A.L., A.B., O.M., M.S.S., M.O-M., A.V.K., M.S.S., P.T.E., C.T.R. and S.Kat.

<u>Functionally-informed fine-mapping</u>: A.G., C.G., M.F., J.C.W., R.C., and H.W. <u>PoPs</u>: E.M.W, H.K.F, K.G.A. and S.Kat.

<u>Mouse knock-outs</u>: P.D.

<u>eQTL data and analysis</u>: A.S.B., T.J., H.S., L.M., J.L.M.B., A.R., and P.D.

<u>Causal gene prioritization</u>: K.G.A., A.S.B., P.S., R.C., A.G., C.G., M.F., J.C.W., and H.W.

<u>Data visualization</u>: C.G., A.G., N.P.B., M.C.C., J.F., D-K.J., A.S.B., and K.G.A.

<u>CARDIoGRAMplusC4D Executive Committee</u>: J.E., N.J.S., H.S., H.W., P.D., R.R., M.F., S.Kat., and J.D.

<u>Conceptualization, initiation and oversight</u>: A.S.B., S.Kat, J.D., C.R., N.J.S., H.S., J.E., H.W., P.D., C.J.W.

<u>Drafted and edited the manuscript</u>: K.G.A., T.J., A.G., S.Kan., B.N.W., J.D., C.T.R., H.K.F., J.C.H., R.C., J.E., N.J.S., H.S., H.W., C.J.W., P.D., S.Kat. and A.S.B.

All authors reviewed the manuscript.

## Notes

### Author Declarations

Ethical approval and informed consent were obtained for all participating studies (where necessary). Further details for the de novo studies can be found in the Supplementary Material. Examples of study-specific oversight bodies include: deCODE: The study was approved by the Data Protection Authority of Iceland and the National Bioethics Committee of Iceland (Approvals No. VSNb2015080003-03.01 and VSNb2015030022-03.01 with amendments). EPIC-CVD: The study was approved by the local ethics committees of the participating centres and the Institutional Review Board of the International Agency for Research on Cancer (IARC, Lyon). HUNT: Participation in the HUNT Study is based on informed consent, and the study has been approved by the Data Inspectorate and the Regional Ethics Committee for Medical Research in Norway. UK Biobank: UK Biobank has approval from the North West Multi-centre Research Ethics Committee (MREC), which covers the UK. It also sought the approval in England and Wales from the Patient Information Advisory Group (PIAG) for gaining access to information that would allow it to invite people to participate.

