## Supplementary Text and Figures for "Discovery and systematic characterization of risk variants and genes for coronary artery disease in over a million participants"

### SUPPLEMENTARY MATERIAL

#### Contents


**Supplementary Figures and Legends**.…...2

**Supplementary Table Legends**.…………...10

**Study Descriptions**………………………....17

**Supplementary References**……………….21

**Consortium Members**…………………….…22

#### Supplementary Figures

##### Supplementary Figure 1. Study design.

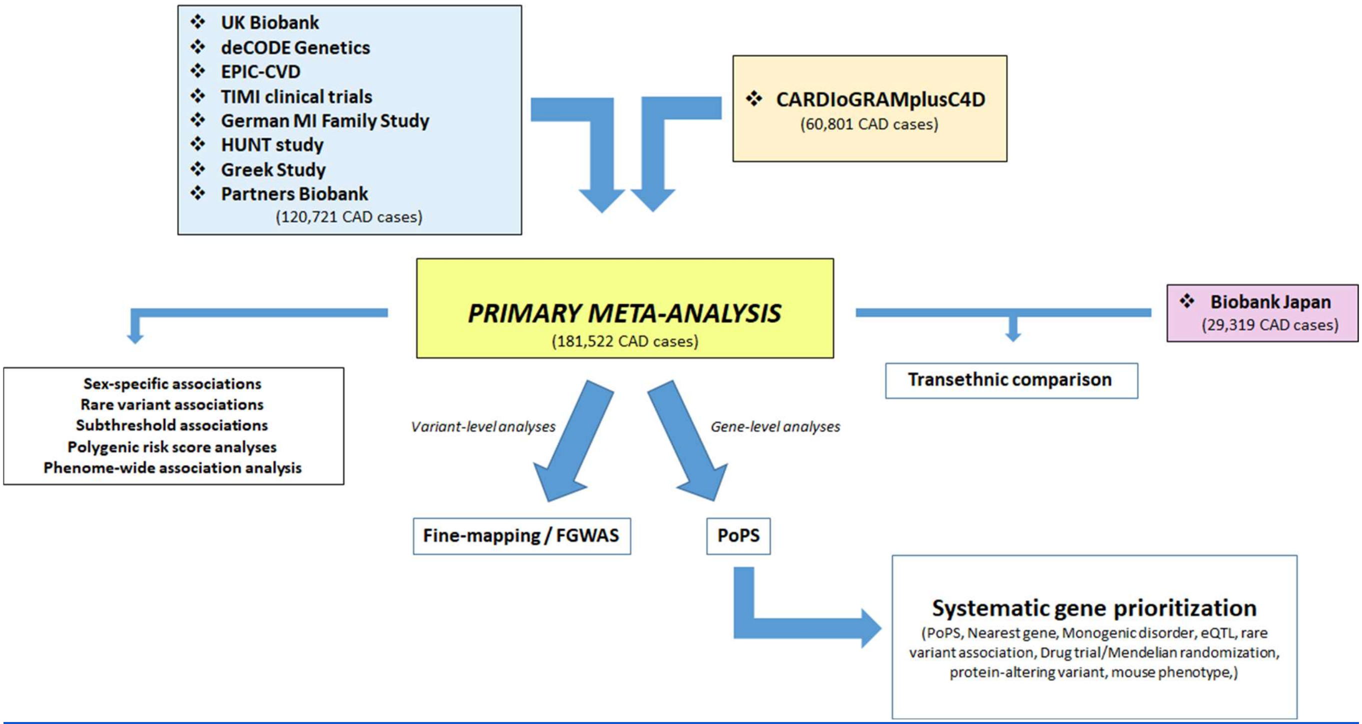

**Supplementary Figure 2. Genetic architecture of 897 association signals for CAD.**

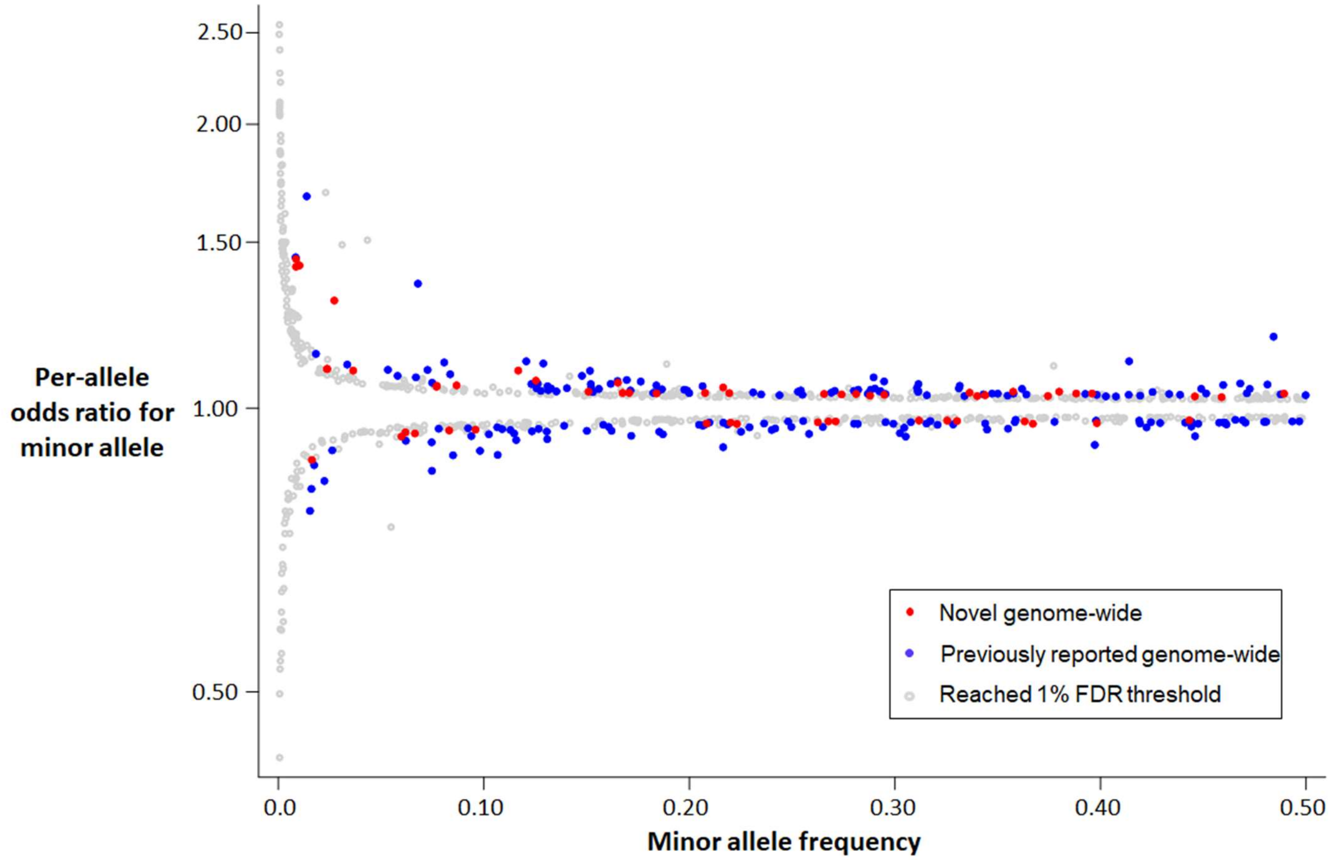


**Minor allele frequency versus per-allele odds ratio for CAD for all sentinel variants reaching genome-wide significance or the 1% FDR threshold in our study**.
Colored circles indicate genome-wide significant associations (p-value<5.0x10$^{-8}$) with sentinel variants that are not correlated ($r^2$<0.2) with a previously reported variant ('novel' – red), genome-wide significant sentinel variants correlated with a previously reported variant ('known' - blue), and associations reaching the 1% FDR threshold (p-value<2.52x10$^{-5}$) in our meta-analysis ('1% FDR' – grey).

**Supplementary Figure 3. Gene-based association testing of rare variants in UK Biobank.**

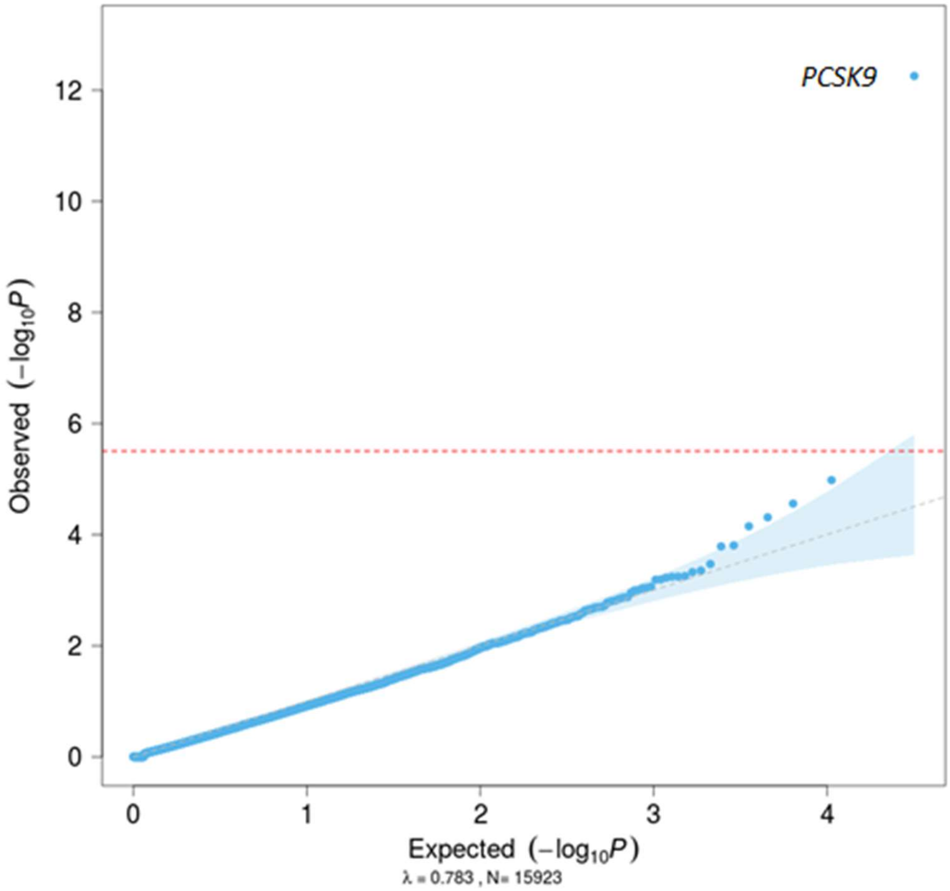


**QQ-plot of aggregate variant association tests from 15,923 genes versus CAD in UK Biobank.** Results presented here are for the SKATO test using the Mask 1 ('lenient') filter, which includes variants with minor allele frequency<5% that are annotated as missense, frameshift, stop gain, stop loss or splice site. Results for all genes, tests and filters are in Supplementary Table 5. Details of masks and tests are in Supplementary Table 4.

**Supplementary Figure 4. Trans-ethnic comparison.**

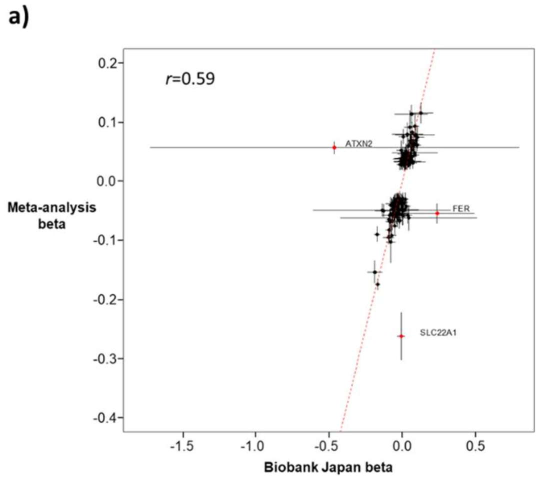


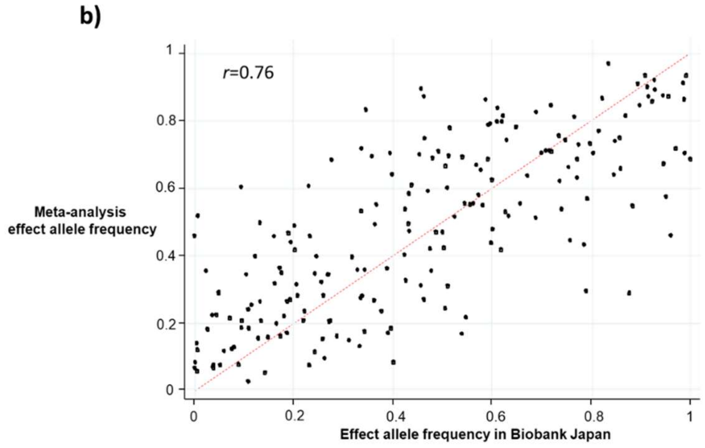


**a) Comparison of beta estimates between the meta-analysis and Biobank Japan.**
Black dots denote the beta estimates for the CAD associations for 199 sentinel variants reaching genome-wide significance in the (predominantly European ancestry) meta-analysis (Y-axis) that were also present in the publicly available summary statistics from Biobank Japan (X-axis). Variants were aligned according to the effect allele in Supplementary Table 2. Horizontal and vertical lines represent 95% confidence intervals. The Pearson correlation coefficient was 0.59, which increased to 0.85 when three outlying variants marked in red (at *ATXN2, FER*, and *SLC22A1*) were excluded.

**b) Comparison of allele frequencies between the meta-analysis and Biobank Japan.**
Black dots denote the allele frequencies for 199 sentinel variants reaching genome-wide significance in the (predominantly European ancestry) meta-analysis (Y-axis) that were also present in the publicly available summary statistics from Biobank Japan (X-axis). Variants were aligned according to the effect allele in Supplementary Table 2. The Pearson correlation coefficient was 0.76.

**Supplementary Figure 5. Epigenetically-informed fine-mapping of the *MAFB* locus.**

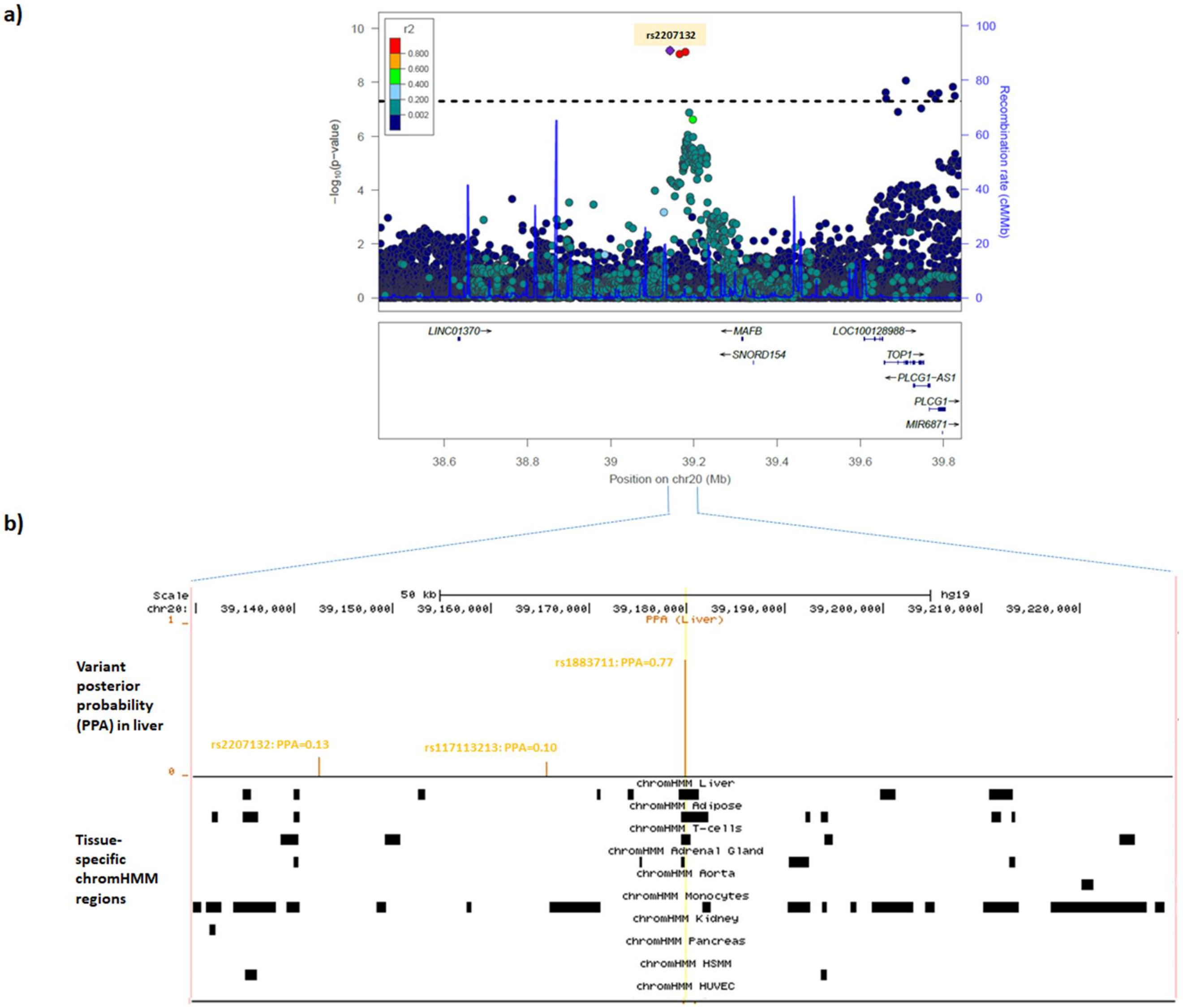


**a) Regional association plot from the CAD meta-analysis for the *MAFB* region.**

Colored dots represent the position (X-axis) in GRCh37 coordinates and –log10(meta-analysis p-value) (Y axis) of each variant in the region. Dots are shaded to represent the $r^2$ with the lead CAD variant (rs2207132), estimated using a random sample of 5,000 European ancestry participants from the UK Biobank. Recombination peaks are plotted in blue based on estimates of recombination from 1000 Genomes European ancestry individuals.

**b) Tissue-specific imputed chromHMM states at the three credible set variants in the *MAFB* region.**

The top track shows the position on chromosome 20 (GRCh37) in the *MAFB* gene region. The second track shows as orange vertical bars the posterior probability (Y-axis) for each variant in the window from the FGWAS fine-mapping, identifying rs1883711 (PPA=0.77) as the most likely causal variant. The third track indicates as a black box the position of the imputed chromHMM state in each of the 10 CAD-relevant

tissues based on epigenomic data from the NIH Roadmap Epigenomics Consortium project. The yellow vertical line indicates the position of the most likely causal variant (rs1883711) with respect to the chromHMM states. rs1883711 lies in an enhancer region for liver (the most strongly enriched tissue for this region) and adipose, the two functionally enriched tissues in the region. The other two variants in the 95% credible set (rs2207132 and rs117113213) do not lie in regions annotated as chromHMM states.
HSMM = human skeletal muscle myoblasts; HUVEC = human umbilical vein endothelial cells; PPA = posterior probability of being the causal variant.

**Supplementary Figure 6. Pairwise concordance of eight gene-prioritization predictors to identify most likely causal genes.**

| | Nearest gene | PoPS | eQTL | Mouse knock-out | Protein-altering variant | Monogenic disorder | Drug/MR | Previous rare variant |
|---|---|---|---|---|---|---|---|---|
| Nearest gene | 149/185 (81%) | 75/185 (41%) | 32/89 (36%) | 24/77 (31%) | 9/41 (22%) | 20/41 (49%) | 8/17 (47%) | 3/10 (30%) |
| PoPS | 108/185 (58%) | 142/185 (77%) | 34/89 (38%) | 21/77 (27%) | 25/41 (61%) | 11/41 (27%) | 5/17 (29%) | 3/10 (30%) |
| eQTL | 53/89 (60%) | 53/89 (60%) | 75/89 (84%) | 11/32 (34%) | 7/18 (39%) | 12/17 (71%) | 1/4 (25%) | 1/2 (50%) |
| Mouse knock-out | 50/77 (65%) | 49/77 (64%) | 20/32 (62%) | 63/77 (82%) | 7/20 (35%) | 6/24 (25%) | 4/10 (40%) | 2/9 (22%) |
| Protein-altering variant | 30/41 (73%) | 15/41 (37%) | 11/18 (61%) | 11/20 (55%) | 33/41 (80%) | 2/7 (29%) | 2/4 (50%) | 1/3 (33%) |
| Monogenic disorder | 18/41 (44%) | 23/41 (56%) | 5/17 (29%) | 16/24 (67%) | 5/7 (71%) | 27/41 (66%) | 3/9 (33%) | 2/7 (29%) |
| Drug/MR | 9/17 (53%) | 9/17 (53%) | 3/4 (75%) | 5/10 (50%) | 2/4 (50%) | 6/9 (67%) | 12/17 (71%) | 0/8 (0%) |
| Previous rare variant | 7/10 (70%) | 6/10 (60%) | 1/2 (50%) | 6/9 (67%) | 2/3 (67%) | 5/7 (71%) | 6/8 (75%) | 8/10 (80%) |

White squares lying on the diagonal contain the number of genes for which that predictor provided evidence (denominator) and the number of times for which that predictor prioritized the most likely causal gene at the locus (numerator). For example, eQTL data provided evidence for 89 causal genes, of which 75 (84%) were also the most likely causal gene at the locus.

Blue squares below the diagonal show the concordance between pairs of predictors and contain the number of genes for which both predictors provided evidence (denominator) and the number of times for which the prioritized causal gene was the same (numerator). For example, the nearest gene and the presence of a protein-altering variant in high LD ($r^2 \geq 0.8$) with the CAD sentinel both provided evidence for a causal gene at 41 loci, of which they were concordant (i.e. prioritized the same causal gene) at 30 (73%). Darker blue squares show higher levels of concordance.

Orange squares above the diagonal show the discordance between pairs of predictors and contain the number of genes for which both predictors provided evidence (denominator) and the number of times for which the prioritized causal gene from either predictor was not the most likely causal gene (numerator). For example, the nearest gene and the presence of a protein-altering variant in high LD ($r^2 \geq 0.8$) with the CAD sentinel both provided evidence for a causal gene at 41 loci, of which they were discordant at 9 (11%). Darker orange squares show higher levels of discordance.

The eight predictors used to prioritize causal genes are:

1) 'Nearest gene' - the nearest gene to the CAD sentinel variant;
2) 'PoPS' - either of the two top prioritized genes in the region from PoPS (Supplementary Table 21);
3) 'eQTL' - a gene in the region has an eQTL in a CAD-relevant tissue from GTEx or STARNET for which the lead eSNP is in high LD ($r^2 \geq 0.8$) with the CAD sentinel variant (Supplementary Tables 24 & 25);
4) 'Mouse knock-out' - a gene for which a mouse knock-out has a cardiovascular-relevant phenotype (Supplementary Table 32);
5) 'Protein-altering variant' - A gene in the region harbors a protein-altering variant that is in high LD ($r^2 \geq 0.8$) with the CAD sentinel variant (Supplementary Table 28);
6) 'Monogenic disorder' - a gene in the region harbors a variant that ClinVar classifies as having evidence for being pathogenic for a cardiovascular-relevant monogenic disorder (Supplementary Table 31);
7) 'Drug/MR' - a gene in the region has been implicated by an effective drug targeting the protein and/or a positive Mendelian randomization (MR) study suggesting a causal effect of the protein on CAD (Supplementary Table 28);
8) 'Previous rare variant' - A gene in the region has been shown to have a rare variant association with CAD in a previous whole-exome sequencing (WES) or genotyping study (Supplementary Table 28).

#### Supplementary Tables

##### Supplementary Table 1. Participant characteristics from contributing studies.

Summary of participant characteristics, genotyping and imputation methods and genetic QC for the ten studies included in the primary CAD GWAS.
AF = atrial fibrillation; HRC = Haplotype Reference Consortium; HWE = Hardy-Weinberg Equilibrium; MAF = minor allele frequency; MI = myocardial infarction; NCSP = Nomesco Classification of Surgical Procedures.

##### Supplementary Table 2. 241 conditionally independent genome-wide significant associations with CAD.

Single variant and joint effect estimates for the 241 conditionally independent associations with CAD that reached genome-wide significance. Positions are according to GRCh37. Beta estimates are per-allele estimates for the effect allele (EA). Order of the studies in the 'Direction' column is: UK Biobank, CARDIoGRAMplusC4D, EPIC-CVD, GerMIFS5, GerMIFS6, GerMIFS7, deCODE, Greek Coronary Disease cohort, HUNT, Partners Biobank, TIMI. 'Novel_region' denotes whether a variant within a 500Kb window has been previously reported to be associated with CAD at genome-wide significance. 'Novel_association' denotes whether a variant with $r^2$>0.2 has been previously reported to be associated with CAD at genome-wide significance. Where the association is known, the previously reported variant that has the strongest correlation with the meta-analysis sentinel variant is listed, along with the pairwise correlation ($r^2$).
EA = effect allele; LCI = lower 95% confidence interval; SE = standard error; UCI = upper 95% confidence interval.

##### Supplementary Table 3. Results from phenome-wide association study (PheWAS) in UK Biobank.

Significant associations with diseases and continuous traits in UK Biobank are listed for each of the 241 conditionally independent genome-wide associations with CAD. Details of the diseases analyzed are provided in Supplementary Table 29 and details of the continuous traits are provided in Supplementary Table 30. Variant positions are given according to GRCh37.

##### Supplementary Table 4. Framework for rare variant tests in UK Biobank.

Details of minor allele frequency thresholds and variant annotations used for each of the four masks used in the rare variant testing. Annotations were generated using the LOFTEE plugin from the Ensembl Variant Effect Predictor.

**Supplementary Table 5. Rare variant test results**

P-values for each combination of gene, test and mask. Results are ordered by ascending P-value for Mask1 SKATO. ‘NA’ denotes that this gene could not be tested using this mask due to insufficient variants. Details of tests and masks are listed in Supplementary Table 4.

**Supplementary Table 6. Studies contributing to the sex analyses.**

Study names and numbers of cases and controls stratified by sex.

**Supplementary Table 7. Variants with significant evidence of sex differences.**

Ten associations reaching genome-wide significance in the sex-differentiation test. Results are provided separately for males and females, as well as in the combined meta-analysis.

**Supplementary Table 8. 897 conditionally independent associations with CAD significant at 1% false-discovery rate.**

Single variant and joint effect estimates for the 897 conditionally independent associations with CAD from among the 47,622 variants that reached the 1% FDR threshold. Positions are according to GRCh37. Beta estimates are per-allele estimates for the effect allele (EA).

**Supplementary Table 9. Additional loci identified through inclusion of recently published summary statistics from Biobank Japan.**

Summary statistics of the sentinel variants for 38 associations that reached genome-wide significance after combining our meta-analysis summary statistics with results from a recently published CAD GWAS from Biobank Japan. Effect allele frequencies, beta estimates and p-values from our meta-analysis, Biobank Japan, and the combined meta-analysis are presented. All variants except the two denoted with an asterisk (rs5867305 and rs75655731) reached the 1% FDR threshold in our meta-analysis.

**Supplementary Table 10. Baseline characteristics of polygenic risk score training and testing datasets from the Malmo Diet and Cancer Study.**

Conventional CAD risk factors and medication usage are summarized for incident CAD cases and non-cases in the PRS training dataset and PRS testing dataset from the Malmo Diet and Cancer Study. Values are mean ± SD unless otherwise stated.
*P-values were obtained from Cox proportional hazard models adjusted for age and sex.

**Supplementary Table 11. Polygenic risk score training - CARDIoGRAMplusC4D-2015, LDpred.**

Details of the performance of 136 LDpred polygenic risk scores in the PRS training dataset from the Malmö Diet and Cancer Study based on the 2015 GWAS summary statistics. Score performance per mean HR or AUC from 100 bootstrapped samples; top performing scores in bold.
HR = Hazard ratio per SD increase in polygenic risk score; AUC = Area under the receiver operating characteristic curve.

**Supplementary Table 12. Polygenic risk score training - CARDIoGRAMplusC4D-2015, Pruning and Thresholding.**

Details of the performance of 45 Pruning and Thresholding polygenic risk scores in the PRS training dataset from the Malmö Diet and Cancer Study based on the 2015 GWAS summary statistics. Score performance per mean HR or AUC from 100 bootstrapped samples; top performing scores in bold.
HR = Hazard ratio per SD increase in polygenic risk score; AUC = Area under the receiver operating characteristic curve.

**Supplementary Table 13. Polygenic risk score training - Current Study, LDpred.**

Details of the performance of 136 LDpred polygenic risk scores in the PRS training dataset from the Malmo Diet and Cancer Study based on the GWAS summary statistics from the current study. Score performance per mean HR or AUC from 100 bootstrapped samples; top performing scores in bold.
HR = Hazard ratio per SD increase in polygenic risk score; AUC = Area under the receiver operating characteristic curve.

**Supplementary Table 14. Polygenic risk score training - Current Study, Pruning and Thresholding.**

Details of the performance of 45 Pruning and Thresholding polygenic risk scores in the PRS training dataset from the Malmo Diet and Cancer Study based on the GWAS summary statistics from the current study. Score performance per mean HR or AUC from 100 bootstrapped samples; top performing scores in bold.
HR = Hazard ratio per SD increase in polygenic risk score; AUC = Area under the receiver operating characteristic curve.

**Supplementary Table 15. Statistical comparisons of best performing polygenic risk scores derived from CARDIoGRAMplusC4D-2015 and Current Study.**

Details of the performance of the best performing polygenic risk scores from each score derivation method and set of GWAS summary statistics in the testing dataset from the Malmö Diet and Cancer Study. Score performance per mean HR or AUC from 100 bootstrapped samples. Score performances compared by Wilcoxon rank-sum test (P-value).
HR = Hazard Ratio per SD increase in the PRS; AUC = Area under the receiver operator characteristic curve.

**Supplementary Table 16. Protein-altering variants at genome-wide significant associations.**

Details of the 44 associations for which the sentinel variant or a strong proxy ($r^2 \geq 0.8$) was a protein-altering variant according to the Ensembl Variant Effect Predictor. $r^2$ estimates are between the sentinel CAD variant and the proxy protein-altering variant(s).
EA_freq = effect allele frequency.

**Supplementary Table 17. Enrichment of chromatin states in ten cardiovascular relevant tissues.**

Fold enrichment (log2) values for the ten tissues that were enriched for chromatin states in the genome-wide meta-analysis summary statistics.

**Supplementary Table 18. Functionally-informed fine-mapping of 116 enriched regions.**

Summary of the tissue-specific enrichment and functionally-informed fine-mapped for the 116 regions enriched in at least one of the ten enriched CAD-relevant tissues. Enrichments showing a >3SD increment in Bayes Factor (BF) are highlighted in red.
NULLBF = Bayes Factor under the null model; BESTBFJUMP = maximum increment (i.e. enrichment) observed across the ten tissues; PPA = posterior probability of association; HUVEC = human umbilical vein endothelial cells; HSMM = human skeletal muscle myoblasts.

**Supplementary Table 19. 95% credible sets for 116 fine-mapped CAD regions.**

Variants included among the 95% credible set (i.e. total PPA>0.95) for each of the 116 fine-mapped regions. Variants are ordered by locus and then in descending order by PPA. Variants with a PPA>0.5 are highlighted in red.

**Supplementary Table 20. Likely causal variants and eQTLs at 49 fine-mapped regions.**

Details of the 49 most likely causal variants with PPA>0.50 including overlap ($r^2 \geq 0.8$) with eQTLs from CAD-relevant tissues from the STARNET and GTEx studies.
eQTL = expression quantitative trait locus.

**Supplementary Table 21. Polygenic Priority Score (PoPS) results - 500kb window.**

PoPS score and locus-specific rank for all genes within a 500kb window of each of the 241 sentinel genome-wide significant associations with CAD; and the top ten PoPS features contributing to the prioritization of each gene.
ENSGID = Ensembl gene ID; Score = raw PoPS score; prioritized = gene with highest PoPS score within 500kb of sentinel.

**Supplementary Table 22. Polygenic Priority Score (PoPS) results - 1MB window.**

PoPS score and locus-specific rank for all genes within a 1MB window of each of the 241 sentinel genome-wide significant associations with CAD; and the top ten PoPS features contributing to the prioritization of each gene.
ENSGID = Ensembl gene ID; Score = raw PoPS score; prioritized = gene with highest PoPS score within 500kb of sentinel.

**Supplementary Table 23. PoPS feature clusters contributing to prioritization of genes for CAD.**

Hierarchical clustering of 21,407 PoPS features into 3,149 clusters, ranked by relative contribution to PoPS scores of genes prioritized for CAD.

**Supplementary Table 24. eQTLs from GTEx in high LD with lead CAD variants.**

eQTL look-ups from CAD-relevant tissues (adipose [subcutaneous, visceral omentum], adrenal gland, artery [aorta, coronary, tibial], liver and whole blood) in GTEx v7. Associations are only shown where the eQTL reached a 5% false-discovery rate threshold and the lead eQTL variant was in high LD ($r^2 \geq 0.8$) with the CAD sentinel variant.
eQTL = expression quantitative trait locus.

**Supplementary Table 25. eQTLs from STARNET in high LD with lead CAD variants.**

eQTL look-ups from CAD-relevant tissues (atherosclerotic aortic root [AOR], atherosclerotic-lesion-free internal mammary artery [MAM], blood [BLD], liver [LIV], subcutaneous fat [SF], skeletal muscle [SKLM], and visceral abdominal fat [VAF]) in STARNET. Associations are only shown where the eQTL reached a 5% false-discovery rate threshold and the lead eQTL variant was in high LD ($r^2 \geq 0.8$) with the CAD sentinel variant.

**Supplementary Table 26. Validation of systematic causal gene prioritization framework using 30 positive control genes for CAD.**

Each row describes the results of the systematic causal gene prioritization framework for one of the 30 a priori defined positive control genes with established causal roles in CAD. The eight predictors used to prioritize causal genes are: the nearest gene to the sentinel variant, presence of a significant rare coding variant association in a previous association study, presence of a pathogenic (or likely) variant for a CAD-relevant monogenic disorder in ClinVar, evidence from Mendelian Randomization or an effective cardiovascular drug, a protein-altering variant in high LD ($r^2 \geq 0.8$) with the sentinel CAD variant, the first- or second-ranked gene by the PoPS method (genes with an asterisk were from a 1Mb window as no gene was prioritized in a 500Kb window around the sentinel variant), an eQTL in a CAD-relevant tissue from GTEx or STARNET in high LD ($r^2 \geq 0.8$) with the CAD sentinel variant, or a cardiovascular-relevant phenotype in a mouse gene knockout for a gene in a CAD-associated region. The 'positive_control_gene' column lists the 30 positive control genes, while the ‘most_likely_causal_gene’ column lists the gene with the maximum number of predictors for any gene within 500Kb of a CAD sentinel variant. Where multiple genes tied for maximum number of predictors, all tied genes are listed.

**Supplementary Table 27. Agreement of 8 predictors for prioritizing causal genes.**

Quantitative summary of the causal gene prioritization framework. For each of the 8 predictors, the agreement of that predictor with the most likely causal gene is provided (numerator), along with the number of times that predictor provided evidence. The top half of the table considers independent genes, whereas the bottom half considers independent associations.

**Supplementary Table 28. Systematic integration of eight predictors to prioritize causal genes at 241 CAD associations.**

Each row describes the results of the systematic causal gene prioritization framework for one of the 241 genome-wide associations with CAD. The eight predictors used to prioritize causal genes are: the nearest gene to the sentinel variant, presence of a significant rare coding variant association in a previous association study, presence of a pathogenic (or likely) variant for a CAD-relevant monogenic disorder in ClinVar, evidence from Mendelian Randomization or an effective cardiovascular drug, a protein-altering variant in high LD ($r^2 \geq 0.8$) with the sentinel CAD variant, the first- or second-ranked gene by the PoPS method (genes with an asterisk were from a 1Mb window as no gene was prioritized in a 500Kb window around the sentinel variant), an eQTL in a CAD-relevant tissue from GTEx or STARNET in high LD ($r^2 \geq 0.8$) with the CAD sentinel variant, or a cardiovascular-relevant phenotype in a mouse gene knockout for a gene in a CAD-associated region. The ‘most_likely_causal_gene’ column lists the gene with the maximum number of predictors for any gene within 500Kb of a CAD sentinel variant. For associations where no gene had more than one supporting predictor, no gene is prioritized. Where multiple genes tied for maximum number of predictors, all tied genes are listed. The table also shows the results of the functionally-informed fine-mapping in FGWAS and the results of the PheWAS in UK Biobank.

**Supplementary Table 29. UK Biobank phenome-wide association scan (PheWAS), phenotypic definitions for clinical diseases.**

Definition and number of cases for each of the 56 disease outcomes tested in the PheWAS in UK Biobank.

**Supplementary Table 30. UK Biobank phenome-wide association scan (PheWAS), list of continuous variables.**

Participant numbers included in analyses of 32 continuous biomarkers or traits in the UK Biobank PheWAS.

**Supplementary Table 31. Genes harboring variants pathogenic for cardiovascular-relevant phenotypes in ClinVar.**

List of genes in CAD-associated regions for which pathogenic (or likely pathogenic) variants were found in ClinVar for phenotypes and disorders related to CAD.

**Supplementary Table 32. Mouse gene knockouts with cardiovascular relevant phenotypes.**

List of cardiovascular-relevant mouse phenotypes for gene knockouts of genes in genome-wide significant association regions. Columns describe the gene (including HGNC and Mouse Genome Informatics IDs), the mouse model ID and description, and the mouse phenotyping ID and description. The table is ordered alphabetically by gene name.

#### STUDY DESCRIPTIONS

EPIC-CVD

EPIC-CVD is a case-cohort study nested within the European Prospective Investigation into Cancer and Nutrition (EPIC) cohort (1). The EPIC cohort consists of 366,521 women and 153,457 men, aged between 35 and 70 years at baseline, recruited from the population at 23 centres across 10 European countries (Denmark, France, Germany, Greece, Italy, The Netherlands, Norway, Spain, Sweden and the UK) between 1992 and 2000. More than 93% of participants were of European ancestry. For EPIC-CVD, a representative subcohort of 17,634 participants was selected by simple random sampling, stratified by centre, from participants who had available stored blood and buffy coat samples (n=385,747) (2, 3).

For this GWAS analysis, prevalent or incident coronary artery disease cases were compared with non-cases from the subcohort. Coronary artery disease was defined as myocardial infarction, chronic ischaemic heart disease or angina using ICD9 (410-414) or ICD10 (I20-I25) codes. Methods used in the recruitment centres to determine first non-fatal CHD and stroke events included self-report and linkage with morbidity or hospital registries. For most centres, non-fatal events were further validated by a review of medical records and/or linkage with registries. Fatal events were generally determined through mortality registries (3).

EPIC complies with the Declaration of Helsinki, and all participants gave written informed consent before participating in this study. The study was approved by the local ethics committees of the participating centres and the Institutional Review Board of the International Agency for Research on Cancer (IARC, Lyon). A list of the EPIC-CVD Principal Investigators can be found at the end of this Supplement. To avoid overlapping participants, EPIC-CVD was removed from the meta-analyses based on the CardioMetabochip and Exome array.

deCODE

The aim of deCODE genetics in Reykjavik, Iceland, is to find associations between variations in the sequence of the genome and human phenotypes. Subjects with a broad range of phenotypes, their relatives, and control subjects have been recruited continuously since 1996 through a variety of research programs, including cardiovascular (4). We assigned coronary artery disease case status based on the relevant ICD-9 and ICD-10 codes and NCSP procedure codes from Landspitali – The National University Hospital, the only tertiary hospital in Iceland, and from the Causes of Death Register. Case status is updated annually. The control group consisted of individuals free of coronary artery disease.

The study was approved by the Data Protection Authority of Iceland and the National Bioethics Committee of Iceland (Approvals No. VSNb2015080003-03.01 and VSNb2015030022-03.01 with amendments). All participants donating samples signed informed consents. Personal identities of those contributing phenotypes and biological samples were encrypted with a third party system, provided by the Data Protection Authority of Iceland. To avoid overlapping participants, deCODE was removed from the meta-analysis based on the CardioMetabochip array.

GerMIFS

The GerMIFS V cases consist of patients referred for coronary angiography, classified as MI cases, from Germany (5). Control samples were recruited as part of the Cooperative Health Research in the Augsburg Region, a population-based study to assess the health status of the population in Augsburg and the surrounding area. The GerMIFS VI cases consist of patients referred for coronary angiography, classified as CAD or MI cases based on the coronary angiogram, with a stenosis diameter of more than 50% in at least one coronary vessel from Germany (6). Control samples consist of patients referred for coronary angiography with exclusion of coronary artery disease and no history of atrial fibrillation. The GerMIFS VII cases consist of patients referred for coronary angiography, classified as CAD or MI cases, with a stenosis diameter of more than 50% in at least one coronary vessel, with a disease manifestation at young age, multivessel disease, history of previous MI/coronary artery bypass graft surgery and/or absence of traditional risk factors from Germany. Control samples were recruited as CAD-free individuals of the Heinz-Nixdorf-Recall Study which recruited samples in the area of Essen, Northrhine-Westfalia of a prospective cardio-vascular focused population-based cohort. The studies were approved by the local ethics committee and participants gave written informed consent before participating in this study, and the studies were conducted in accordance with the Declaration of Helsinki.

Greek Coronary Disease Cohort (GCC)

GCC is a case-control study conducted in Greece. CAD cases included participants hospitalized for acute coronary syndrome or diagnosed with Left Main CAD during hospitalization. Controls were drawn from the TEENs of Attica: Gene and Environment study (TEENAGE) (7) and from the Non-alcoholic Fatty Liver Disease study (NAFLD) case-control study. The GCC study had obtained local ethics approval and all patients had provided written informed consent prior to enrolment.

HUNT study

The Nord-Trøndelag Health Study (HUNT) is a population-based health survey conducted in the county of Nord-Trøndelag, Norway, where the entire county's population aged 20 years or older were invited to participate (8). Four waves of the survey have been conducted, HUNT1 [1984-1986], HUNT2 [1995-1997], HUNT3 [2006-2008], and HUNT4 [2017-2019], enrolling more than 123,000 individuals at one or more HUNT surveys with participation rates of 89%, 70%, 54%, and 54% respectively. We used a combination of hospital, out-patient, and emergency room discharge diagnoses (ICD-9 and ICD-10) to identify 7,710 CAD cases and 58,577 controls with genotype data. The 'intermediate CAD' phenotype was defined in HUNT as myocardial infarction (I21-I24, 410), chronic IHD (I25.1, I25.2, I25.5, I25.6, I25.7, I25.8, I25.9, 411, 412, 414.0, 414.8, 414.9), or self-reported history of CABG, which was used in the absence of ICD codes for CABG, triple heart bypass or PTCA. Angina cases (I20, 413) were excluded from controls. As a prospective study, we identify prevalent and incident cases. The genotyped cohort has 32748 males and 36887 females. Participation in the HUNT Study is based on informed consent, and the study has been approved by the Data Inspectorate and the Regional Ethics Committee for Medical Research in Norway. To avoid overlapping participants, HUNT was removed from the meta-analysis based on the Exome array.

Partners Biobank

The Partners Healthcare Biobank is a large research data and sample repository comprising more than 100,000 participants that is embedded within the framework of Partners Personalized Medicine (9). Participants are prospectively enrolled in the context of outpatient visits, inpatient stays, and emergency department encounters. The Partners Biobank contains banked samples (plasma, serum, DNA and buffy coats), genomic data, and other health information, including data from the electronic health record (EHR) at hospitals affiliated with the Partners Healthcare system – primarily the Massachusetts General Hospital and the Brigham and Women's Hospital. Array-based genotyping was performed using either the Illumina Multi-Ethnic Genotyping Array, Expanded Multi-Ethnic Genotyping Array, or the Multi-Ethnic Global BeadChip Array (Illumina, Inc., San Diego, CA). We studied the first 13,667 genotyped participants from the Partners Biobank with relevant clinical data available.

TIMI

Three cardiovascular outcomes trials from the TIMI Study Group were included in the GWAS analysis: PEGASUS-TIMI 54, ENGAGE AF-TIMI 48, and SAVOR-TIMI 53. These trials contributed a combined 26,737 patients (17,887 cases and 8,850 controls).

The PEGASUS-TIMI 54 trial was a multinational, randomized, double-blind, placebo-controlled trial of the efficacy of ticagrelor among patients with prior MI (10). The inclusion criteria were age of at least 50 years old with one additional high-risk feature: 65 years or older, diabetes, a second MI, multivessel coronary artery disease, or renal dysfunction. Exclusion criteria included known bleeding disorder, gastrointestinal bleeding within 6 months, history of ischemic stroke, intracranial bleeding, central nervous system abnormality, surgery within 30 days, or planned use of $P2Y_{12}$ receptor antagonist, dipyridamole, cilostazol, or an anticoagulant. Patients were randomized 1:1:1 to either ticagrelor 90 mg twice daily, 60 mg twice daily, or placebo and followed for a median of 2.8 years. The mean age of the study population was 65 years old and 76% were men. Comorbidities included smoking (17%), hypertension (78%), diabetes (32%), prior PCI (83%), and prior CABG (60%). All 10,607 patients who consented for genetic analysis, passed QC, and were of European ancestry were included in this GWAS analysis.

ENGAGE AF-TIMI 48 was a 3-arm multinational, randomized, double-blind, placebo-controlled trial comparing two doses of the Xa inhibitor edoxaban to warfarin in patients with atrial fibrillation (11). The inclusion criteria were age of at least 21 years old with atrial fibrillation, a $CHADS_2$ score of 2 or higher, and on anticoagulation. Exclusion criteria included atrial fibrillation with reversible etiology, kidney dysfunction, high risk of bleeding, use of DAPT, significant mitral stenosis, or recent cardiovascular events. Patients were randomized 1:1:1 to either warfarin, low dose edoxaban, or high dose edoxaban and followed for a median of 2.8 years. The mean age of the study population was 72 years old and 62% were men. Comorbidities included diabetes (38%), stroke (28%), and heart failure (57%). All 9,647 patients who consented for genetic analysis, passed QC, and were of European ancestry were included in this GWAS analysis.

The SAVOR-TIMI 53 trial was a multinational, randomized, double-blind, placebo-controlled trial of the DPP4 inhibitor saxagliptin in diabetics (12). The inclusion criteria were age of at least 40 years with type 2 diabetes mellitus, a glycated hemoglobin of 6.5% to 12.0%, and either established cardiovascular disease or multiple cardiovascular risk factors. Key exclusion criteria included end-stage renal disease, kidney transplantation, or incretin-based therapy within 6 months. Patients were randomized 1:1 to receive saxagliptin or placebo and followed for a median of 2.9 years. The mean age of the study population was 65 years old and 67% were men. Comorbidities included atherosclerosis (78%) and HTN (81%). All 6,483 patients who consented for genetic analysis, passed QC, and were of European ancestry were included in this GWAS analysis.

UK Biobank

UK Biobank (UKB) is a prospective study of approximately 500,000 individuals comprising members of the UK population aged 40-69 collected from multiple sites across the United Kingdom from 2006-2010 (14). For GWAS we utilized version 3 of the UK Biobank genotype data which was imputed to 1000 Genomes, UK10K, and Haplotype Reference Consortium panels (doi: https://doi.org/10.1038/s41586-018-0579-z, 10.1038/ng.3679) for 472,335 individuals of European ancestry that had not withdrawn consent for study as of April, 2018. 33,941 cases were defined as prevalent or incident MI or intervention (CABG, PTCA) ascertained from Hospital Episode Statistics or self-report; or IHD death ascertained from Office of National Statistic. 438,394 controls were taken from the remaining population considered to be CAD and angina free. The minimum MAF for testing was $1 \times 10^{-6}$.

#### CONSORTIUM MEMBERS

1) **CARDIoGRAMplusC4D Consortium and collaborators**

Majid Nikpay, Anuj Goel, Hong-Hee Won, Leanne M Hall, Stavroula Kanoni, Danish Saleheen, Theodosios Kyriakou, Christopher P Nelson, Jemma C Hopewell, Thomas R Webb, Abbas Dehghan, Maris Alver, Sebastian M Armasu, Kirsi Auro, Andrew Bjonnes, Daniel I Chasman, Shufeng Chen, Ian Ford, Nora Franceschini, Christian Gieger, Christopher Grace, Stefan Gustafsson, Jie Huang, Shih-Jen Hwang, Yun Kyoung Kim, Marcus E Kleber, King Wai Lau, Xiangfeng Lu, Yingchang Lu, Leo-Pekka Lyytikäinen, Evelin Mihailov, Alanna C Morrison, Natalia Pervjakova, Liming Qu, Lynda M Rose, Elias Salfati, Richa Saxena, Markus Scholz, Albert V Smith, Emmi Tikkanen, Andre Uitterlinden, Xueli Yang, Weihua Zhang, Wei Zhao, Mariza de Andrade, Paul S de Vries, Natalie R van Zuydam, Sonia S Anand, Lars Bertram, Frank Beutner, George Dedoussis, Philippe Frossard, Dominique Gauguier, Alison H Goodall, Omri Gottesman, Marc Haber, Bok-Ghee Han, Jianfeng Huang, Shapour Jalilzadeh, Thorsten Kessler, Inke R König, Lars Lannfelt, Wolfgang Lieb, Lars Lind, Cecilia M Lindgren, Marja-Liisa Lokki, Patrik K Magnusson, Nadeem H Mallick, Narinder Mehra, Thomas Meitinger, Fazal-Ur-Rehman Memon, Andrew P Morris, Markku S Nieminen, Nancy L Pedersen, Annette Peters, Loukianos S Rallidis, Asif Rasheed, Maria Samuel, Svati H Shah, Juha Sinisalo, Kathleen E Stirrups, Stella Trompet, Laiyuan Wang, Khan S Zaman, Diego Ardissino, Eric Boerwinkle, Ingrid B Borecki, Erwin P Bottinger, Julie E Buring, John C Chambers, Rory Collins, L Adrienne Cupples, John Danesh, Ilja Demuth, Roberto Elosua, Stephen E Epstein, Tõnu Esko, Mary F Feitosa, Oscar H Franco, Maria Grazia Franzosi, Christopher B Granger, Dongfeng Gu, Vilmundur Gudnason, Alistair S Hall, Anders Hamsten, Tamara B Harris, Stanley L Hazen, Christian Hengstenberg, Albert Hofman, Erik Ingelsson, Carlos Iribarren, J Wouter Jukema, Pekka J Karhunen, Bong-Jo Kim, Jaspal S Kooner, Iftikhar J Kullo, Terho Lehtimäki, Ruth J F Loos, Olle Melander, Andres Metspalu, Winfried März, Colin N Palmer, Markus Perola, Thomas Quertermous, Daniel J Rader, Paul M Ridker, Samuli Ripatti, Robert Roberts, Veikko Salomaa, Dharambir K Sanghera, Stephen M Schwartz, Udo Seedorf, Alexandre F Stewart, David J Stott, Joachim Thiery, Pierre A Zalloua, Christopher J O'Donnell, Muredach P Reilly, Themistocles L Assimes, John R Thompson, Jeanette Erdmann, Robert Clarke, Hugh Watkins, Sekar Kathiresan, Ruth McPherson, Panos Deloukas, Heribert Schunkert, Nilesh J Samani, Martin Farrall.

2) **EPIC-CVD Consortium**

Principal investigators of the EPIC-CVD Consortium include the following:

Kim Overvad, Christina Dahm, Anne Tjønneland, Marie-Christine Boutron, Rudolf Kaaks, Matthias Schulze, Pietro Ferrari, Giovanna Masala, Vittorio Krogh, Salvatore Panico, Rosario Tumino, Giuseppe Matullo, Carlotta Sacerdote, Jolanda Boer, Yvonne van der Schouw, Guri Skeie, J. Ramon Quiros, María-José Sánchez Pérez, Conchi Moreno-Iribas, María Dolores

Chirlaque, Pilar Amiano, Liher Imaz, Olle Melander, Patrik Wennberg, Nicholas J. Wareham, Timothy J. Key, Elisabete Weiderpass, Elio Riboli, Adam S. Butterworth and John Danesh.

The coordination of EPIC is financially supported by International Agency for Research on Cancer (IARC) and also by the Department of Epidemiology and Biostatistics, School of Public Health, Imperial College London which has additional infrastructure support provided by the NIHR Imperial Biomedical Research Centre (BRC). The national cohorts are supported by: Danish Cancer Society (Denmark); Ligue Contre le Cancer, Institut Gustave Roussy, Mutuelle Générale de l'Education Nationale, Institut National de la Santé et de la Recherche Médicale (INSERM) (France), German Cancer Aid, German Cancer Research Center (DKFZ), German Institute of Human Nutrition Potsdam-Rehbruecke (DIfE), Federal Ministry of Education and Research (BMBF) (Germany); Associazione Italiana per la Ricerca sul Cancro-AIRC-Italy, Compagnia di SanPaolo and National Research Council (Italy); Dutch Ministry of Public Health, Welfare and Sports (VWS), Netherlands Cancer Registry (NKR), LK Research Funds, Dutch Prevention Funds, Dutch ZON (Zorg Onderzoek Nederland), World Cancer Research Fund (WCRF), Statistics Netherlands (The Netherlands); Health Research Fund (FIS) - Instituto de Salud Carlos III (ISCIII), Regional Governments of Andalucía, Asturias, Basque Country, Murcia and Navarra, and the Catalan Institute of Oncology - ICO (Spain); Swedish Cancer Society, Swedish Research Council and County Councils of Skåne and Västerbotten (Sweden); Cancer Research UK (14136 to EPIC-Norfolk; C8221/A29017 to EPIC-Oxford), Medical Research Council (1000143 to EPIC-Norfolk; MR/M012190/1 to EPIC-Oxford) (United Kingdom).

We thank all EPIC participants and staff for their contribution to the study, the laboratory teams at the Medical Research Council Epidemiology Unit for sample management, Cambridge Genomic Services for genotyping, Matthew Walker and Sarah Spackman (BHF Cardiovascular Epidemiology Unit) for data management and the team at the EPIC-CVD Coordinating Centre for study coordination and administration.
